# Comparative Evaluation of Mosquito Repellent Products in South Asia and North America: Efficacy, Safety, and Public Health Implications

**DOI:** 10.64898/2026.06.07.26355094

**Authors:** Kazi Sahal, S M Al Amin, Tamanna Mostafa, Sibao Wang, Barbara Colucci, Md Ushama Shafoyat, Zhi-ming Yuan, Gong Cheng

## Abstract

Mosquito-borne diseases continue to pose significant public health challenges worldwide, particularly in densely populated regions of South Asia and parts of North America experiencing increasing vector prevalence due to climate and environmental changes. Commercial mosquito repellents are widely used as a primary preventive measure; however, their efficacy, safety, and public health impacts vary depending on formulation, active ingredients, environmental conditions, and user practices. This study presents a comparative evaluation of commonly used mosquito repellent products in South Asia and North America, including coils, vaporizers, sprays, creams, and Natural repellents. The research aims to assess repellent efficacy against major mosquito vectors, evaluate potential health and respiratory effects associated with prolonged exposure, and analyze consumer awareness and usage patterns across different regions. Laboratory-based efficacy testing and field observations were conducted to compare protection duration, repellency rate, and environmental performance under varying climatic conditions. Safety assessments included analysis of chemical composition, indoor air quality impact, and reported adverse health symptoms among users. The findings indicate significant differences in effectiveness and safety profiles among product categories and geographical regions. Synthetic repellents generally demonstrated higher repellency duration, while herbal formulations showed improved safety and environmental compatibility. The study highlights the importance of standardized evaluation protocols, regulatory oversight, and public awareness in promoting safe and effective mosquito control strategies. These findings may support policymakers, healthcare professionals, and manufacturers in improving mosquito repellent technologies and reducing the burden of mosquito-borne diseases globally.

## 1. Introduction

Mosquito-borne diseases remain among the most serious global public health concerns, contributing substantially to morbidity and mortality in tropical and subtropical regions (Peng et al., 2022). Diseases such as dengue fever, malaria, chikungunya, Zika virus infection, and West Nile fever continue to threaten millions of people annually, particularly in densely populated and climatically favorable regions of South Asia and parts of North America (Tuetun et al., 2005). Rapid urbanization, climate change, increased international travel, inadequate sanitation, and the expansion of mosquito habitats have intensified the transmission of vector-borne diseases worldwide. Among the mosquito species responsible for disease transmission, *Aedes aegypti*, *Aedes albopictus*, and *Anopheles* mosquitoes are recognized as major vectors affecting both developing and developed countries (Katz et al., 2008; Kongkaew et al., 2011).

In the absence of universally effective vaccines and complete vector eradication strategies, personal protective measures remain essential for reducing human–mosquito contact. Among these preventive measures, mosquito repellent products are widely used due to their accessibility, affordability, and ease of application (Maia & Moore, 2011; Webb & Hess, 2016). Commercial repellents are available in multiple formulations, including mosquito coils, electric vaporizers, aerosol sprays, topical creams and lotions, and natural or plant-based repellents. These products are designed to repel or incapacitate mosquitoes through chemical or botanical active ingredients such as N,N-Diethyl-meta-toluamide (DEET), pyrethroids, allethrin, citronella oil, eucalyptus oil, neem extract, and lemongrass oil (Nguyen et al., 2023a).

Mosquito coils and electric vaporizers are extensively used in many South Asian countries because of their low cost and effectiveness in indoor environments. However, prolonged exposure to smoke and volatile compounds emitted by these products has raised concerns regarding respiratory irritation, indoor air pollution, and other potential health risks (Nguyen et al., 2023a). Aerosol sprays and topical creams provide rapid, direct protection against mosquito bites, though their effectiveness may vary with concentration, exposure duration, climatic conditions, and user compliance (Chen-Hussey et al., 2014). In recent years, rising public concern about chemical toxicity and environmental sustainability has driven increased interest in natural and herbal repellents, which are often perceived as safer alternatives despite questions about their long-term efficacy and consistency (Nguyen et al., 2023a).

The effectiveness of mosquito repellent products can vary considerably across regions due to differences in mosquito species, environmental conditions, humidity, temperature, housing structures, and consumer behavior. South Asia experiences a high burden of mosquito-borne diseases due to tropical climatic conditions and dense urban populations, leading to widespread daily use of repellent products (Maia & Moore, 2011; Webb & Hess, 2016). In contrast, North America has witnessed the emergence and spread of vector-borne diseases in recent decades, thereby increasing consumer demand for advanced, environmentally friendly mosquito protection technologies. Despite the widespread use of these products, comparative studies evaluating their efficacy, safety profiles, and public health implications across different regions remain limited (Katz et al., 2008).

Furthermore, concerns regarding human exposure to chemical compounds in repellents have become increasingly important. Several studies have reported associations between long-term exposure to mosquito coil smoke and respiratory disorders, headaches, allergic reactions, and indoor particulate pollution (Peterson et al., 2006). Similarly, misuse or overexposure to aerosol sprays and synthetic topical repellents may contribute to dermatological and neurological effects in sensitive populations, including children and pregnant women (Chen-Hussey et al., 2014). Although natural repellents are generally considered safer, scientific evidence regarding their protective duration and effectiveness under real-world conditions remains insufficient.

Therefore, a comprehensive comparative evaluation of mosquito repellent products is necessary to understand their relative performance, safety, and broader public health implications (Maia & Moore, 2011). This study aims to assess and compare commonly used mosquito repellents in South Asia and North America, including coils, vaporizers, sprays, creams, and natural repellents. The research focuses on evaluating repellency effectiveness, duration of protection, potential health and environmental impacts, and consumer usage patterns (Diaz, 2016). The findings of this study are expected to contribute to evidence-based public health recommendations, support regulatory decision-making, and encourage the development of safer, more effective, and environmentally sustainable mosquito repellent technologies (Tamanna et al., 2026).

## 2. Materials and Methods

### 2.1. Study Design

This study was designed as a comparative experimental and observational evaluation of commercially available mosquito repellent products commonly used in South Asia and North America. The investigation focused on determining the efficacy, safety, and public health implications of different repellent formulations, including mosquito coils, electric vaporizers, aerosol sprays, topical creams, and natural repellents. Both laboratory-based and field-based assessments were conducted to evaluate product performance under controlled and real-world environmental conditions(Lee, 2018).

### 2.2. Mosquito Repellent Products

A total of 25 commercially available mosquito repellent products were selected based on market popularity, availability, and formulation type. The products were categorized into five groups:

a. Mosquito Coil: Goodknight Coil, Mortein Coil, Baygon Mosquito Coil, Herbal Mosquito Coil, PIC Mosquito Coil
b. Electric Vaporizer: Goodknight Liquid Vaporizer, All Out Vaporizer, Raid Plug-In, PIC Mosquito Repellent Plug-In, Herbal Vaporizer
c. Aerosol Spray: HIT Mosquito Spray, Mortein PowerGard Spray, OFF! Deep Woods Spray, Cutter Backwoods Spray, Herbal Mosquito Spray
d. Topical Cream/Lotion: Odomos Cream, Goodknight Fabric Roll-On, OFF! FamilyCare Lotion, Sawyer Picaridin Lotion, Herbal Mosquito Cream
e. Natural Repellent: Leeings Mosquito Repellent liquid, Neem Oil Repellent, Lemon Eucalyptus Spray, Lemongrass Repellent, Cedarwood Mosquito Repellent

Products were collected from retail markets and pharmacies in selected countries of South Asia and North America. Product information, including active ingredients, concentration, manufacturer details, and recommended usage instructions, was documented.

### 2.3. Test Mosquitoes

The Foshan (Guangdong, China) strain of Ae. albopictus, collected from Foshan City in 1983, was used in this study(Afify et al., 2019). This mosquito was kept in the laboratory without insecticide exposure. All mosquitoes, including larval and adult stages, were maintained at 28 ± 1 ◦C and 80 ± 5% relative humidity under a 14:10 h (light/dark) photoperiod in the insectary (Carroll et al., 2008).

### 2.4. Laboratory Equipment and Instruments

The following instruments and materials were used: Environmental test chamber, Mosquito exposure cages, Indoor air quality monitor, Particulate matter (PM2.5 and PM10) sensor, Carbon monoxide (CO) detector, Volatile organic compound (VOC) analyzer, Thermo-hygrometer, Stopwatch and digital timer, Protective laboratory equipment, Human skin patch test kits, Statistical analysis software.

### 2.5. Human Volunteers

Six adult volunteers (three males and three females aged 18 to 22 years) participated in the Arm-in-Cage Test Procedure experiments and Dermatological Safety Test. They were non-tobacco users and had no history of dermatosis or allergic reactions to arthropod bites or repellents. Given that various factors may alter a person’s attractiveness to mosquitoes, which may, in turn, affect the results of mosquito repellent tests, all the volunteers were required not to apply any aromatics, skincare, or other repellent products on the skin of the hands 12 h before and during the experiment. The experiment was approved by the Institutional Review Board: Biomedical Research and Innovation Center, Military Institute of Science and Technology, Mirpur -1216, Dhaka, Bangladesh (approval code: 2025H009, Date of Approval: 12/ 09/2025, Clinical trial number: not applicable).

### 2.6. Efficacy Evaluation

#### 2.6.1. Arm-in-Cage Test Procedure

The efficacy comparison of 25 commercial skin-applied repellents against Ae. albopictus females were conducted using the guidelines for the efficacy testing of mosquito repellents for human skin by the WHO. A total of 305 7-day-old starved female Ae. albopictus were placed in a net cage (40 × 30 × 30 cm) before the experiment. The tests were carried out between 15:00 and 20:00 h in accordance with the mosquito biting habits of Ae. albopictus in the 24-h cycle. Volunteers wore long-sleeve white coats to cover their arms, sterilized their hands and wrists with unscented soap, rinsed with water, rinsed with a solution of 70% ethanol or isopropyl alcohol in water, and put on gloves after drying hands with towels. Specific rubber gloves were designed to expose only a 4 × 4 cm area on the back of the hand skin (Figure 1a–e), and the gloves were replaced in each test in case of the repellent residue effect (*Guidelines for Efficacy Testing of Mosquito Repellents for Human Skin*, n.d.).

**Figure 1.**
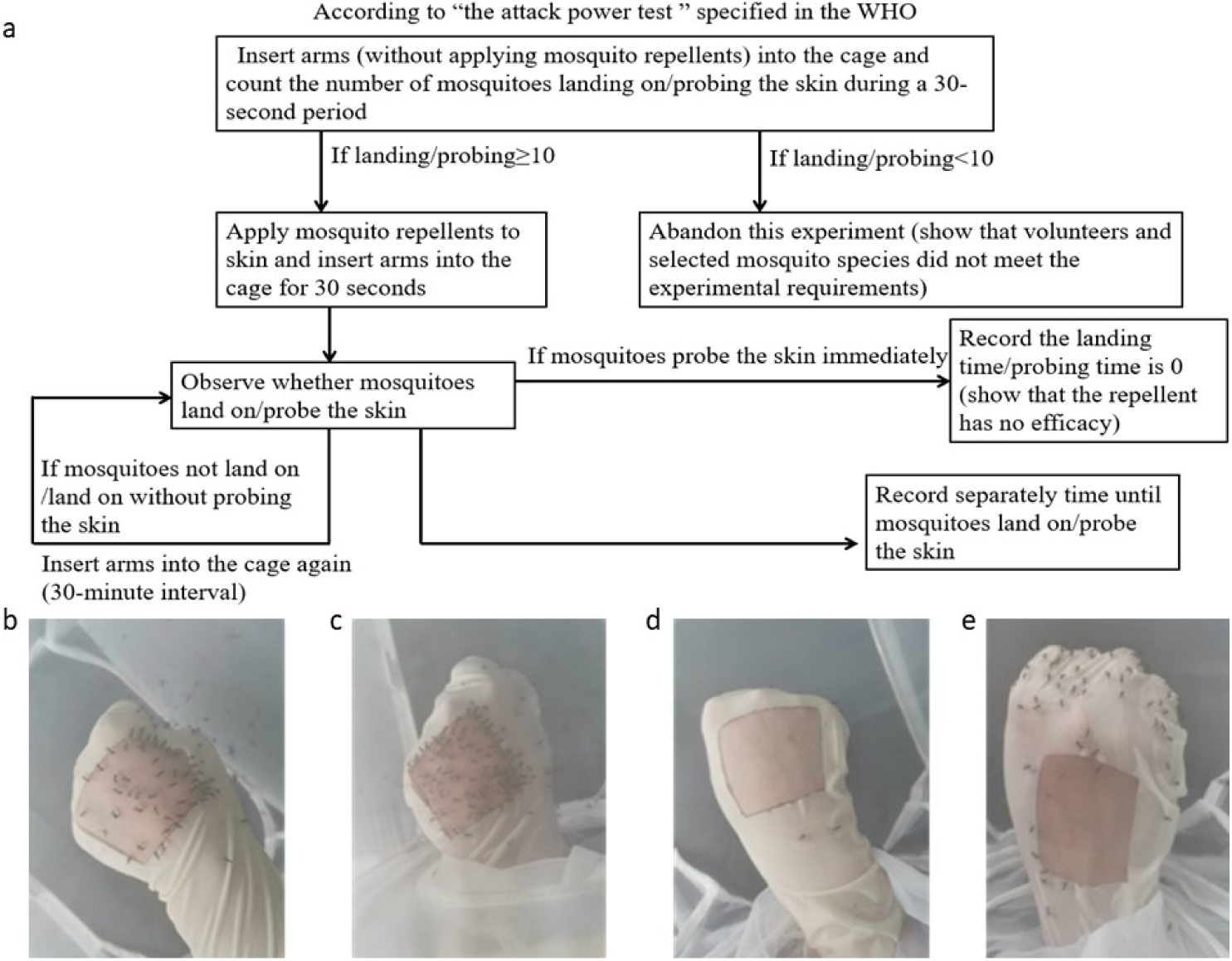
A mosquito attack power testing: (a) pre-experiment design; (b, c) more than 10 mosquitoes landed/probed; (d, e) less than 10 mosquitoes landed/probed. Only if the biting rate was ≥10 landings and/or probings in 30 s, the mosquitoes and volunteers were qualified for further testing.

The first step was to insert the untreated hand into the cage and to count the number of mosquitoes that landed on and/or probed the skin in 30 s. During testing, the volunteer should avoid movement of the hand. Only if the biting rate was ≥10 landings and/or probing in 30 s, the mosquitoes and volunteers were qualified for further testing (Figure 1). The hand was then treated with 50 µL of a repellent product on the exposed 4 × 4 cm skin area, allowed to dry for 1 min, and then inserted into the cage for 30 s (Figure 2). The insertion was repeated at 30-min intervals until the hands received one or more landings and/or probing. Landing and/or probing behavior implies the ending point of the test. A repellent provides efficacy by a reduction in the biting and/or landing activity of mosquitoes. However, landing is not always related to probing, and separate recordings of each behavior are required. The landing and probing times were estimated from the application of the tested products to the mosquitoes’ first biting or landing. Replicate tests repeated this process using different repellents over several weeks. Each repellent had five or six replicate tests conducted per volunteer, allowing the number of results sufficient for statistical analysis. Moreover, volunteers rotated their positions during every test to reduce the potential factors of disturbing the repellent’s efficacy such as positions, catching ability, and the status of mosquitoes. The mosquitoes were allowed a 30-min recovery time after 10 successive tests.

**Figure 2.**
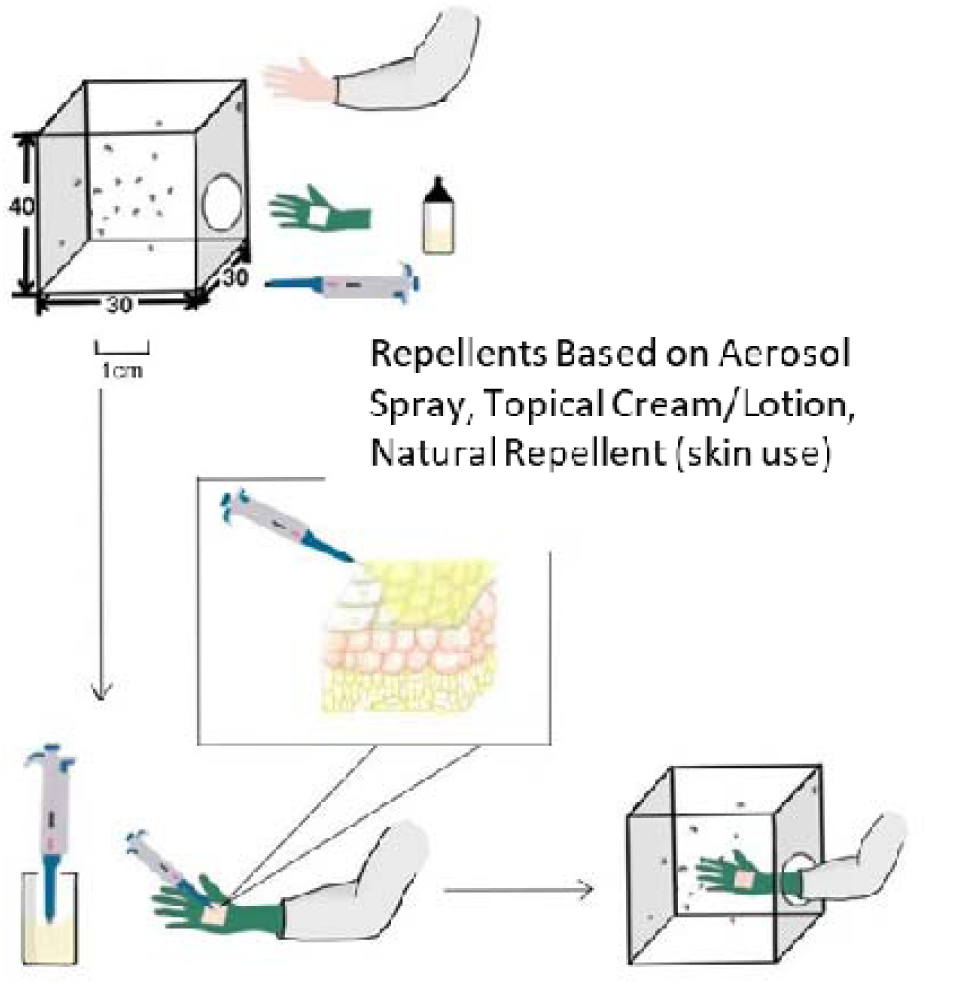
A brief process of efficacy testing of mosquito repellents for human skin in the laboratory.

Repellency percentage was calculated as:

Where:

- C = bites on control arm
- T = bites on treated arm

#### 2.6.2. Room Chamber Test for Coils, Electric Vaporizers, Aerosol spray, Natural Repellent

The efficacy of mosquito coils, electric vaporizers, aerosol sprays, and natural repellents was evaluated using a controlled room chamber test designed to simulate typical residential indoor conditions, as illustrated in Figure 3. The experimental room chamber measured approximately 20 feet × 20 feet × 10 feet and was used to assess the repellency performance of different commercial mosquito repellent products. For each experiment, fifty laboratory-reared female mosquitoes were released into the chamber prior to product exposure. The selected repellent products, including mosquito coils, electric vaporizers, aerosol sprays, and diffuser-based natural repellents, were operated according to the manufacturers’ recommended instructions. Mosquito behavior and activity were continuously monitored for a total duration of 60 minutes. The effectiveness of each product was determined by measuring entomological parameters including knockdown rate, mortality rate, feeding inhibition, and spatial repellency. Observations and data recordings were conducted at specific time intervals of 10, 20, 30, and 60 minutes to evaluate both immediate and prolonged repellent effects under controlled environmental conditions.

**Figure 3.**
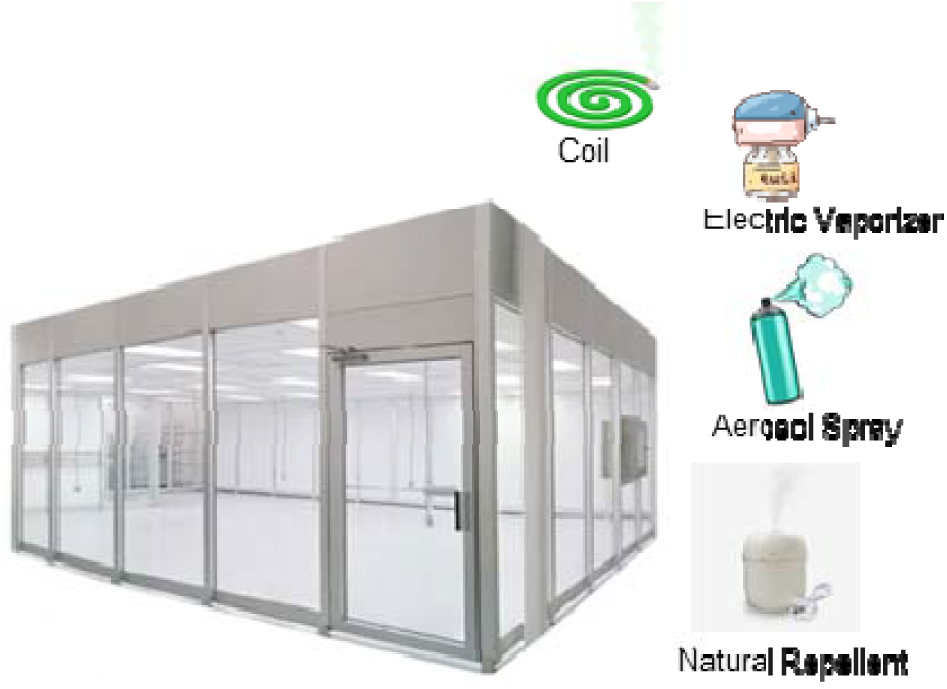
A brief process of efficacy testing of mosquito repellents Room Chamber Test for Coils, Electric Vaporizers, Aerosol spray, Natural Repellent.

#### 2.6.3. Field Evaluation

Field evaluations were conducted in selected residential areas during peak mosquito activity periods (July to October) to assess the real-world performance of mosquito repellent products under natural environmental conditions. Volunteers were assigned different repellent products, including coils, vaporizers, aerosol sprays, topical creams, and natural repellents in 100 households located in selected urban and rural regions of South Asia (Dhaka North City Corporation (DNCC) and Savar Upazila Pathalia Union, Bangladesh) and North America (Los Angeles, California and The Florida Everglades / Southern Florida), and instructed to use them during evening hours when mosquito activity was highest. During the testing period, mosquito landing counts on exposed body areas were recorded to determine the level of protection provided by each product. In addition to efficacy assessment, user comfort, convenience, odor acceptability, and overall satisfaction were evaluated through structured questionnaires completed by the participants after product use. Environmental conditions during field testing were also monitored, including ambient temperature, relative humidity, wind speed, and local mosquito density, as these factors could influence mosquito behavior and repellent effectiveness. The collected data were used to compare product performance and user acceptability under practical residential conditions.

### 2.7. Safety Assessment

#### 2.7.1. Indoor Air Quality Analysis

Indoor air quality analysis was conducted to evaluate the pollutants generated by mosquito coils, electric vaporizers, aerosol sprays, and Natural Repelents (Leeings Mosquito repellent liquid) when used in enclosed residential environments. The experiments were performed in controlled indoor rooms to simulate typical household conditions during repellent use. Air quality parameters monitored during the study included particulate matter concentrations (PM2.5 and PM10), carbon monoxide (CO) levels, and volatile organic compounds (VOCs), which are commonly associated with combustion- and chemical-based repellent products. Measurements were recorded at three different stages: before product activation to establish baseline indoor air conditions, during active product exposure to assess immediate pollutant generation, and after one hour of continuous operation to determine the extent of pollutant accumulation over time. The collected data were used to compare the environmental safety profiles of different mosquito repellent products and to assess their potential impacts on indoor air quality and human respiratory health (Mapossa et al., 2020).

#### 2.7.2. Dermatological Safety Test

The dermatological safety of topical mosquito repellent products, including creams, lotions, sprays, and herbal formulations, was evaluated using a standard skin patch testing method to determine their potential for causing skin irritation or allergic reactions (Sutthanont et al., 2026). Small quantities of each repellent product were carefully applied to designated skin patch areas on volunteer participants of 20 under controlled conditions. The treated skin areas were then monitored continuously for a period of 24 to 48 hours to observe any adverse dermatological responses following product exposure. Particular attention was given to identifying common skin reactions such as redness, itching, irritation, and allergic responses. The severity and frequency of these reactions were documented and compared among different product formulations to assess their relative dermatological safety and suitability for regular human use (Islam et al., 2017).

#### 2.7.3. Respiratory and User Symptom Survey

A structured questionnaire-based survey was conducted among 56 regular users of mosquito repellent products to identify potential health-related symptoms associated with prolonged exposure to coils, vaporizers, aerosol sprays, topical repellents, and natural formulations. Participants were asked about the frequency, duration, and type of repellent products used in their households, along with any physical discomfort experienced during or after product exposure. The survey specifically evaluated commonly reported symptoms, including headache, eye irritation, coughing, breathing discomfort, skin irritation, and nausea(Izadi et al., 2017). Respondents were also asked to indicate the frequency and severity of these symptoms using standardized response scales. The collected information was analyzed to assess the relationship between repellent exposure and possible respiratory or general health effects, thereby contributing to the overall safety evaluation and public health implications of commonly used mosquito repellent products.

### 2.8. Toxicological Evaluation on Animal model

An in vivo toxicological assessment was conducted using a rat model to evaluate the potential health effects of commonly used mosquito repellent products (Peterson et al., 2006). The study focused on representative formulations, including mosquito coil emissions, liquid vaporizer and aerosol exposure, DEET-based topical application, and natural repellent formulations (Chen-Hussey et al., 2014; Magesh et al., 2018; Webb & Hess, 2016). The experimental protocol was designed to simulate realistic exposure scenarios relevant to human use patterns.

#### 2.8.1. Experimental Animals

Healthy adult Wistar rats (*Rattus norvegicus*), weighing 180–220 g, were used for the study. Animals were housed under controlled laboratory conditions (temperature: 22 ± 2°C; relative humidity: 50–60%; 12-hour light/dark cycle) with free access to standard feed and water. All experimental procedures were conducted in accordance with institutional ethical guidelines and approved by the Ethical Committee (Approval Number: 2025H005; Date: 20 Jun 2025).

#### 2.8.2. Grouping and Exposure Protocol

Animals were randomly divided into five groups (n = 3 per group):

- **Group I** (Control): No exposure
- **Group II** (Coil Exposure): Exposure to mosquito coil smoke (8 hours/day)
- **Group III (**Vaporizer and aerosol Exposure): Exposure to vaporizer and aerosol emissions (8 hours/day)
- **Group IV** (Skin Application): Topical application of Topical Cream/Lotion (standard dermal dose)
- **Group V** (Natural Repellent): Exposure using a humidifier (8 hours/day)

The exposure duration was maintained for 28 consecutive days to assess sub-chronic toxicity.

#### 2.8.3. Ethical Considerations, Anesthesia, and Humane Endpoints

All animal experiments were conducted in accordance with institutional ethical guidelines and internationally accepted principles for the care and use of laboratory animals.

##### (a) Methods of sacrifice

At the end of the experimental period, animals were humanely sacrificed under deep anesthesia to minimize pain and distress. Euthanasia was performed by overdose administration of ketamine–xylazine followed by cervical dislocation to ensure rapid and irreversible death prior to tissue collection. This method was selected in accordance with standard laboratory animal welfare recommendations.

##### (b) Methods of anesthesia and/or analgesia

For procedures involving handling and sample collection, animals were anesthetized using an intraperitoneal injection of Ketamine (80–100 mg/kg body weight) combined with Xylazine (5–10 mg/kg body weight) to ensure adequate anesthesia, analgesia, and muscle relaxation during experimental procedures. The depth of anesthesia was monitored through assessment of pedal withdrawal and corneal reflexes prior to invasive manipulations.

##### (c) Efforts to alleviate suffering

Throughout the study, several measures were implemented to alleviate animal suffering, including daily health monitoring, maintenance of appropriate housing conditions, provision of adequate nutrition and hydration, and minimization of unnecessary handling. Animals exhibiting severe distress, persistent respiratory difficulty, excessive weight loss, or signs of severe pain were designated for immediate humane intervention or euthanasia according to predefined humane endpoint criteria.

#### 2.8.4. Toxicological Assessment Parameters

To evaluate potential systemic and organ-specific toxicity associated with mosquito repellent exposure, a comprehensive assessment was conducted in experimental animals, covering clinical, physiological, hematological, biochemical, and histopathological parameters (Fradin, 1998). Animals were observed daily for general health status, behavioral changes, and physiological signs such as altered activity, grooming behavior, lethargy, respiratory distress (dyspnea, nasal irritation, and abnormal breathing patterns), skin irritation, and changes in food and water intake, which served as early indicators of acute or sub-chronic toxic effects. Body weight was recorded weekly to monitor growth and overall health, and at termination, lungs, liver, and kidneys were excised, blotted, and weighed to calculate organ-to-body weight ratios (organ indices), indicating possible organ enlargement, atrophy, or physiological stress due to exposure. Hematological analysis of blood samples included hemoglobin (Hb), total leukocyte count (TLC), and differential leukocyte count (DLC) to evaluate immune response, inflammation, and potential disturbances in blood-forming systems. Biochemical evaluation of serum ALT and AST assessed hepatic function, while creatinine and blood urea levels were used to determine renal function, with deviations indicating possible hepatotoxicity and nephrotoxicity. Finally, histopathological examination of lung, liver, and skin tissues, processed with formalin fixation and Hematoxylin and Eosin (H&E) staining, was performed to detect inflammatory infiltration, cellular degeneration, necrosis, and structural or architectural changes, providing definitive evidence of tissue damage and corroborating clinical and biochemical findings for a comprehensive toxicity assessment (Fradin, 1998).

### 2.9. Public Health and Consumer Awareness Assessment

A cross-sectional public health and consumer awareness survey was conducted among 100 households located in selected urban and rural regions of South Asia (Dhaka North City Corporation (DNCC) and Savar Upazila Pathalia Union, Bangladesh) and North America (Los Angeles, California and The Florida Everglades / Southern Florida) to evaluate patterns of mosquito repellent use and public understanding of mosquito-borne disease prevention. Structured questionnaires were distributed to participants to collect information regarding the frequency of repellent use, preferred product types, awareness of product safety, and knowledge of mosquito-borne diseases, user satisfaction, and perceptions of product affordability. The survey aimed to assess both behavioral and socioeconomic factors influencing the selection and regular use of mosquito repellent products in different geographical settings. The collected data were analyzed to better understand public awareness, safety perceptions, and the broader public health implications associated with mosquito repellent use across diverse populations.

### 2.10. Statistical Analysis

Experimental data obtained from laboratory tests, field evaluations, safety assessments, and consumer surveys were analyzed using appropriate statistical software packages. Descriptive statistics were used to summarize the data and present the overall distribution of measured variables. Comparative analyses among different mosquito repellent products and study groups were performed using one-way analysis of variance (ANOVA) and independent-samples t-tests, as appropriate. The Chi-square test was applied to evaluate associations between categorical variables, particularly in survey-based public health and consumer awareness assessments. Pearson correlation analysis was also conducted to examine relationships between environmental factors, repellent efficacy, and reported health effects. All results were expressed as mean ± standard deviation (SD). Statistical significance was determined at a confidence level of p < 0.05, which was considered indicative of a statistically significant difference between the study variables.

## 3. Result and Discussion

### 3.1. Selected Repellent Products

Finally, we purchased the 25 repellent products widely available in market. The 25 selected repellents were classified into five kinds regarding the active ingredients (Figure 4 a–e). All information on the manufacturer, properties, main components, concentrations, and the toxicity of mosquito repellents is available in Table 3. Also same type of active ingredients products available in market which are not included in our test have same toxicity effect.

**Figure 4.**
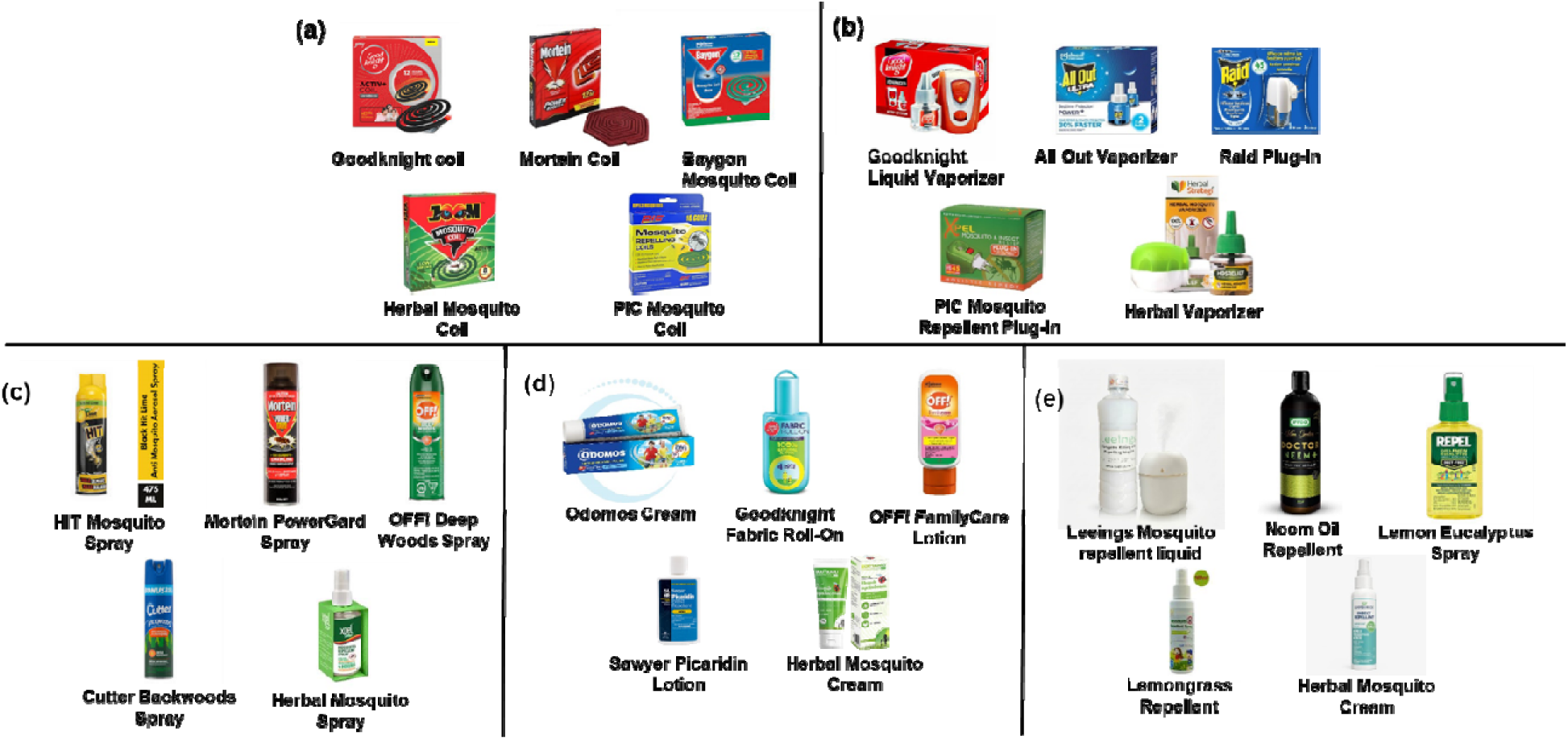
The commercial repellent products selected for testing (25 in total). (a) Mosquito coil; (b) Electric Vaporizer; (c) Aerosol Spray; (d) Topical Cream/Lotion; (e) Natural Repellent.

### 3.2. Comparative Performance Analysis

The comparative evaluation of mosquito repellent products demonstrated considerable variation in efficacy, protection duration, safety profile, and user acceptability among different formulation types used in South Asia and North America. Synthetic repellents containing DEET, picaridin, pyrethroids, transfluthrin, and prallethrin generally showed higher repellency effectiveness and longer protection duration compared with herbal and natural formulations (figure 5). Among the evaluated products, electric vaporizers: Raid Plug-In, and Goodknight Liquid Vaporizer provided the most sustained protection 8-9 hour against *Ae. albopictus* mosquito bites under both laboratory and field conditions. Products such as topical lotions and: OFF! FamilyCare Lotion, Sawyer Picaridin Lotion, exhibited high repellency rates and prolonged protection times exceeding 7-8 hours in most trials. Mosquito coils and aerosol sprays showed rapid knockdown and strong short-term repellency effects 5-6 hour; however, their efficacy gradually decreased with prolonged exposure periods. In Natural repellents Leeings mosquito repellent liquid exhibited high repellency rates and prolonged protection times exceeding 6-8 hours in most trials which is better than all other natural mosquito repellents. Regional differences were also observed between South Asian and North American products. South Asian consumers relied more heavily on coils and vaporizers because of affordability and widespread availability, whereas North American consumers preferred topical sprays and lotions for personal outdoor protection. Overall, the findings indicate that although synthetic repellents provide superior protection efficiency, natural repellents may offer safer alternatives for sensitive populations and environmentally conscious users.

**Figure 5:**
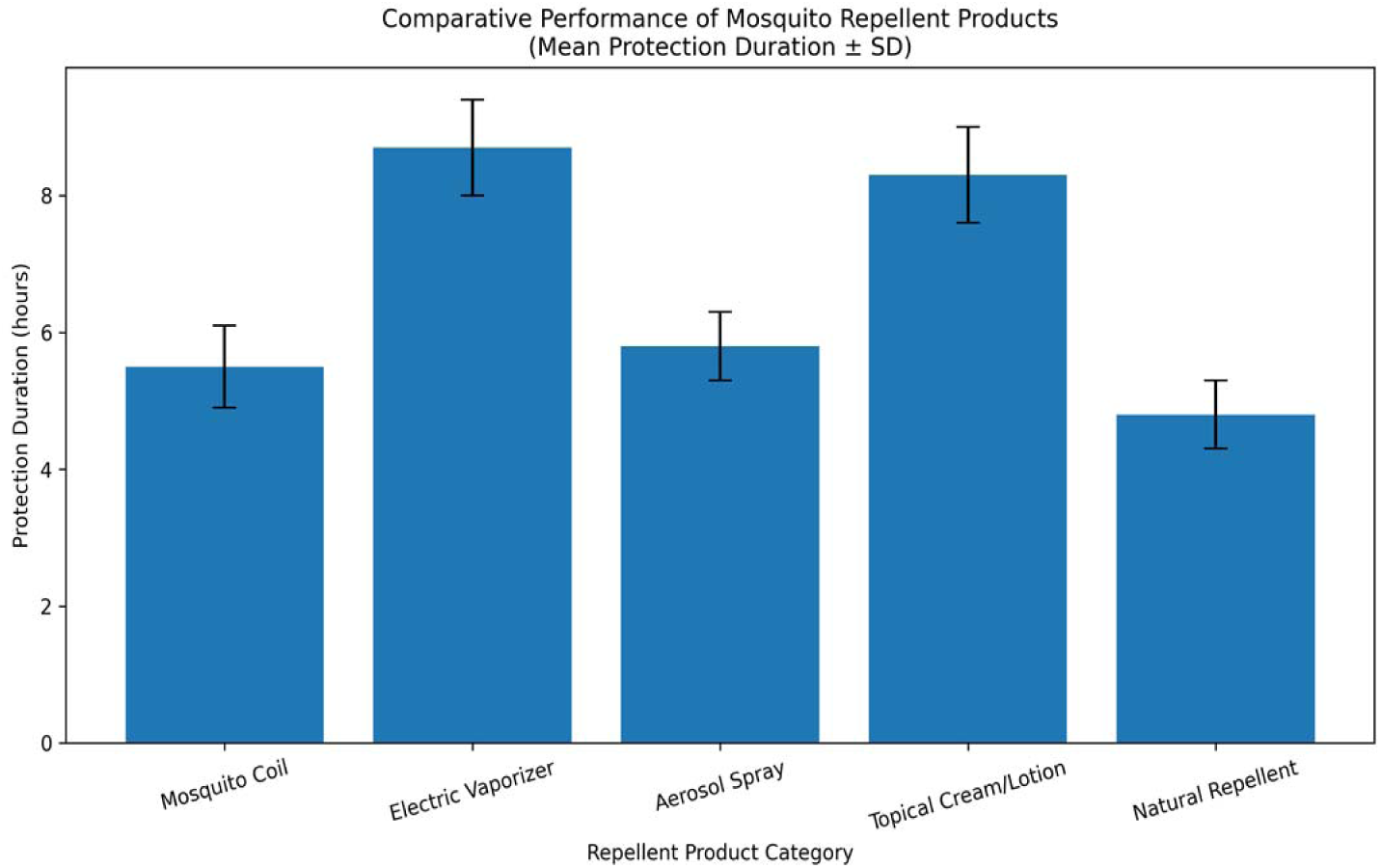
Comparative performance of mosquito repellent products demonstrated protection duration hours among different formulation types used in South Asia and North America.

#### 3.2.1. Arm-in-Cage Repellency Bioassay

The Arm-in-Cage repellency bioassay demonstrated that all tested mosquito repellent products significantly reduced mosquito landing and probing behavior compared with untreated controls (table 1). Topical formulations containing picaridin and DEET exhibited the highest levels of protection against *Ae. albopictus*. Sawyer Picaridin Lotion and OFF! FamilyCare Lotion showed the longest mean landing and probing protection times, while Odomos Cream and OFF! Deep Woods Spray also provided effective repellency for more than 6 hours.

**Table 1.** Protection time of commercial mosquito repellents against *Ae. albopictus* bites.

| Repellent Products | Landing Time (Mean ± SD, h) | Landing Time Range (h) | Probing Time (Mean ± SD, h) | Probing Time Range (h) | Number of Subjects |
| --- | --- | --- | --- | --- | --- |
| Odomos Cream | 7.0 ± 0.5 | 6–8 | 6.5 ± 0.4 | 6–7 | 5 |
| Goodknight Fabric Roll-On | 6.8 ± 0.5 | 6–8 | 6.2 ± 0.4 | 5–7 | 5 |
| OFF! FamilyCare Lotion | 9.2 ± 0.7 | 8–10 | 8.6 ± 0.5 | 8–9 | 5 |
| Sawyer Picaridin Lotion | 10.1 ± 0.8 | 9–12 | 9.4 ± 0.6 | 8–11 | 5 |
| Herbal Mosquito Cream | 4.5 ± 0.5 | 4–5 | 4.0 ± 0.4 | 3–5 | 6 |
| Neem Oil Repellent | 4.8 ± 0.4 | 4–5 | 4.2 ± 0.3 | 3–5 | 5 |
| Lemon Eucalyptus Spray | 5.9 ± 0.5 | 5–6 | 5.3 ± 0.4 | 4–6 | 5 |
| Lemongrass Repellent | 3.5 ± 0.4 | 3–4 | 3.0 ± 0.3 | 2–4 | 6 |
| Cedarwood Mosquito | 4.2 ± 0.5 | 3–5 | 3.8 ± 0.4 | 3–4 | 6 |
| Repellent |  |  |  |  |  |

#### 3.2.2. Room chamber Test result for Coils, Electric Vaporizers, Aerosol spray, Natural Repellent

The room chamber experiments demonstrated that mosquito coils, vaporizers, aerosol sprays, and natural repellents produced varying levels of mosquito knockdown, mortality, and spatial repellency (table 2). Electric vaporizers achieved the highest sustained spatial repellency over the 60-minute observation period, while aerosol sprays produced rapid initial knockdown effects within the first 20 minutes of exposure. Mosquito coils showed effective mortality rates 80-minute but also generated high particulate emissions during combustion. Natural repellents produced average mosquito mortality rates. Natural repellent Leeings mosquito repellent liquid demonstrated repellency over the 70-minute without substantial indoor air pollution. Products containing eucalyptus and citronella oils showed improved spatial repellency compared with other herbal formulations. Overall, synthetic pyrethroid-based products demonstrated significantly higher efficacy than botanical repellents under controlled chamber conditions (p < 0.05).

**Table 2.**
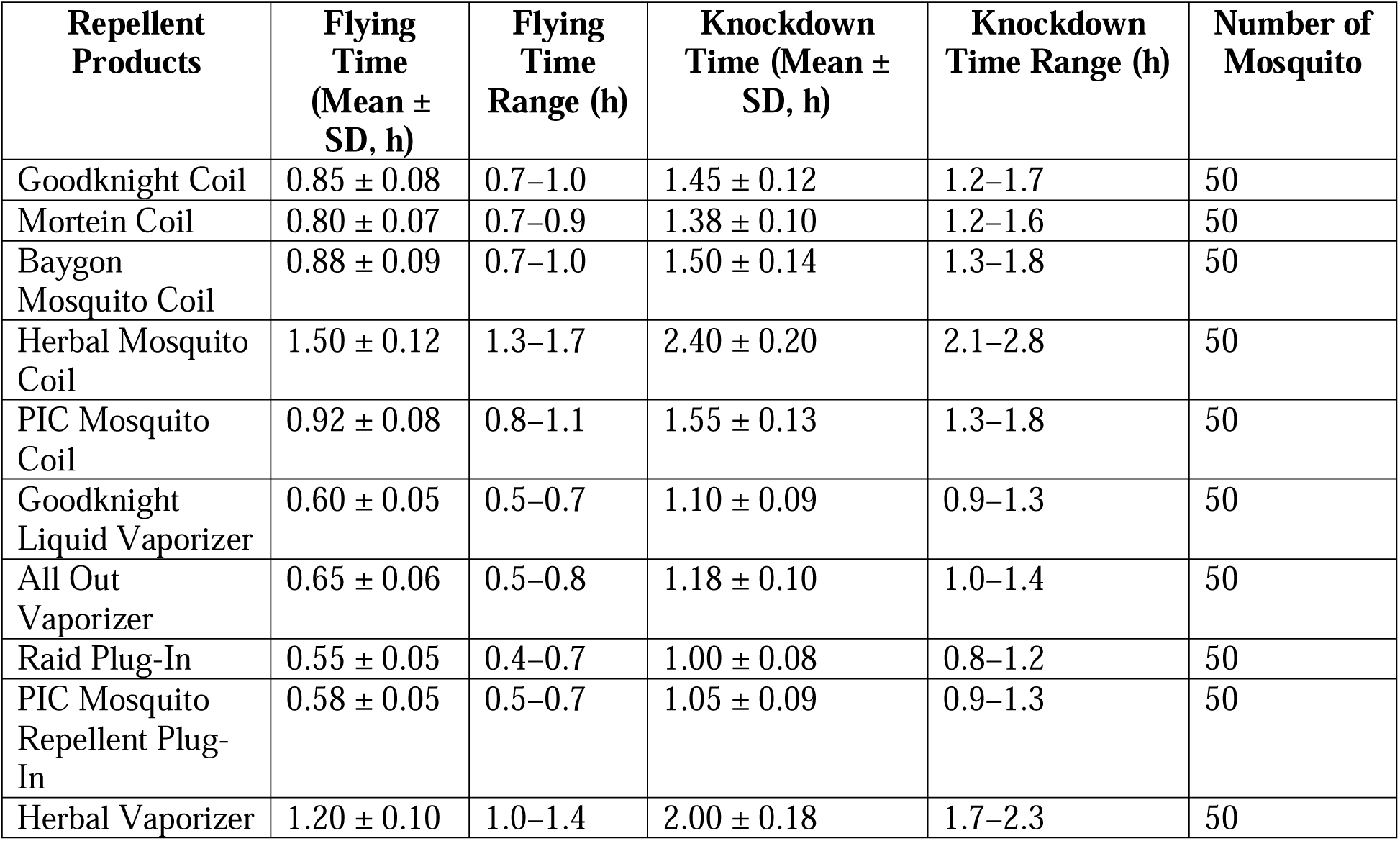
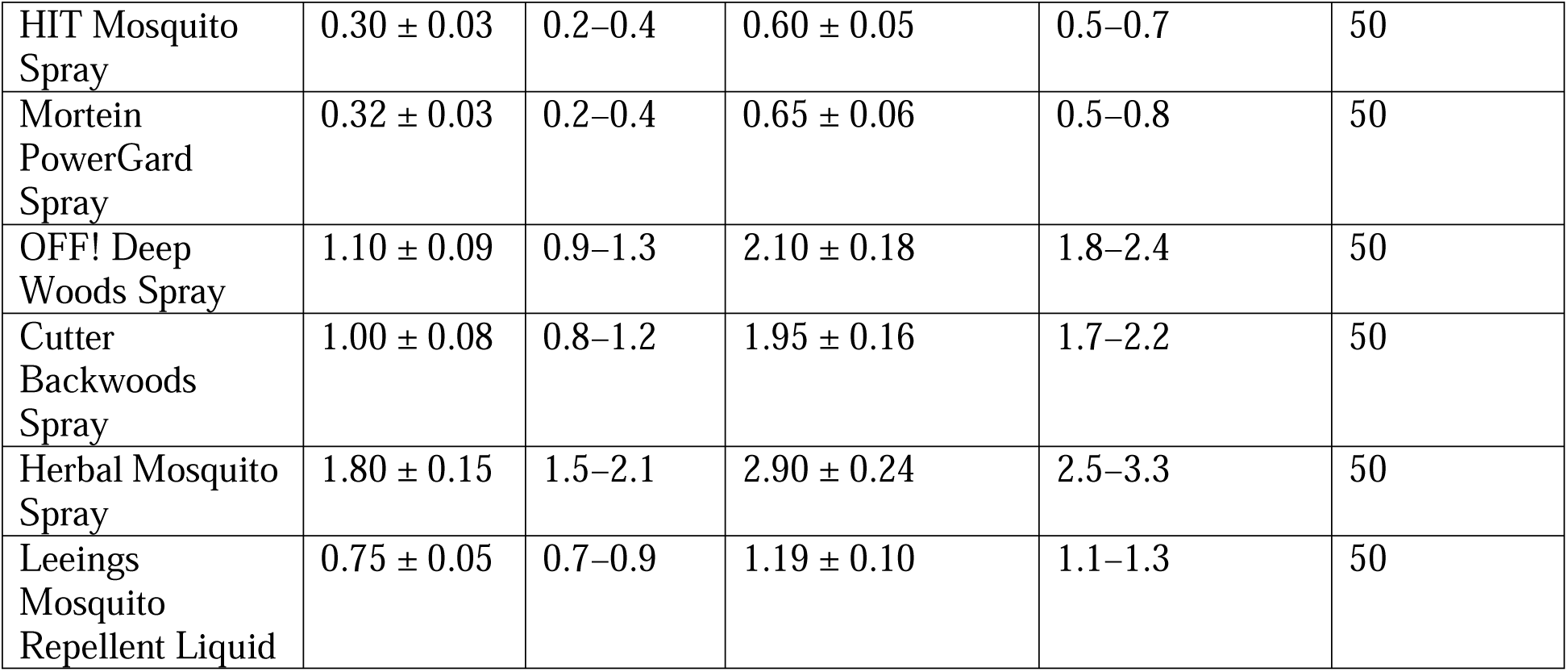
Knockdown time of commercial mosquito repellents against *Ae. albopictus*.

| Repellent Products | Flying Time (Mean ± SD, h) | Flying Time Range (h) | Knockdown Time (Mean ± SD, h) | Knockdown Time Range (h) | Number of Mosquito |
| --- | --- | --- | --- | --- | --- |
| Goodknight Coil | 0.85 ± 0.08 | 0.7–1.0 | 1.45 ± 0.12 | 1.2–1.7 | 50 |
| Mortein Coil | 0.80 ± 0.07 | 0.7–0.9 | 1.38 ± 0.10 | 1.2–1.6 | 50 |
| Baygon Mosquito Coil | 0.88 ± 0.09 | 0.7–1.0 | 1.50 ± 0.14 | 1.3–1.8 | 50 |
| Herbal Mosquito Coil | 1.50 ± 0.12 | 1.3–1.7 | 2.40 ± 0.20 | 2.1–2.8 | 50 |
| PIC Mosquito Coil | 0.92 ± 0.08 | 0.8–1.1 | 1.55 ± 0.13 | 1.3–1.8 | 50 |
| Goodknight Liquid Vaporizer | 0.60 ± 0.05 | 0.5–0.7 | 1.10 ± 0.09 | 0.9–1.3 | 50 |
| All Out Vaporizer | 0.65 ± 0.06 | 0.5–0.8 | 1.18 ± 0.10 | 1.0–1.4 | 50 |
| Raid Plug-In | 0.55 ± 0.05 | 0.4–0.7 | 1.00 ± 0.08 | 0.8–1.2 | 50 |
| PIC Mosquito Repellent Plug-In | 0.58 ± 0.05 | 0.5–0.7 | 1.05 ± 0.09 | 0.9–1.3 | 50 |
| Herbal Vaporizer | 1.20 ± 0.10 | 1.0–1.4 | 2.00 ± 0.18 | 1.7–2.3 | 50 |
| HIT Mosquito Spray | $0.30 \pm 0.03$ | 0.2–0.4 | $0.60 \pm 0.05$ | 0.5–0.7 | 50 |
| Mortein PowerGard Spray | $0.32 \pm 0.03$ | 0.2–0.4 | $0.65 \pm 0.06$ | 0.5–0.8 | 50 |
| OFF! Deep Woods Spray | $1.10 \pm 0.09$ | 0.9–1.3 | $2.10 \pm 0.18$ | 1.8–2.4 | 50 |
| Cutter Backwoods Spray | $1.00 \pm 0.08$ | 0.8–1.2 | $1.95 \pm 0.16$ | 1.7–2.2 | 50 |
| Herbal Mosquito Spray | $1.80 \pm 0.15$ | 1.5–2.1 | $2.90 \pm 0.24$ | 2.5–3.3 | 50 |
| Leeings Mosquito Repellent Liquid | $0.75 \pm 0.05$ | 0.7–0.9 | $1.19 \pm 0.10$ | 1.1–1.3 | 50 |

**Table 3.**
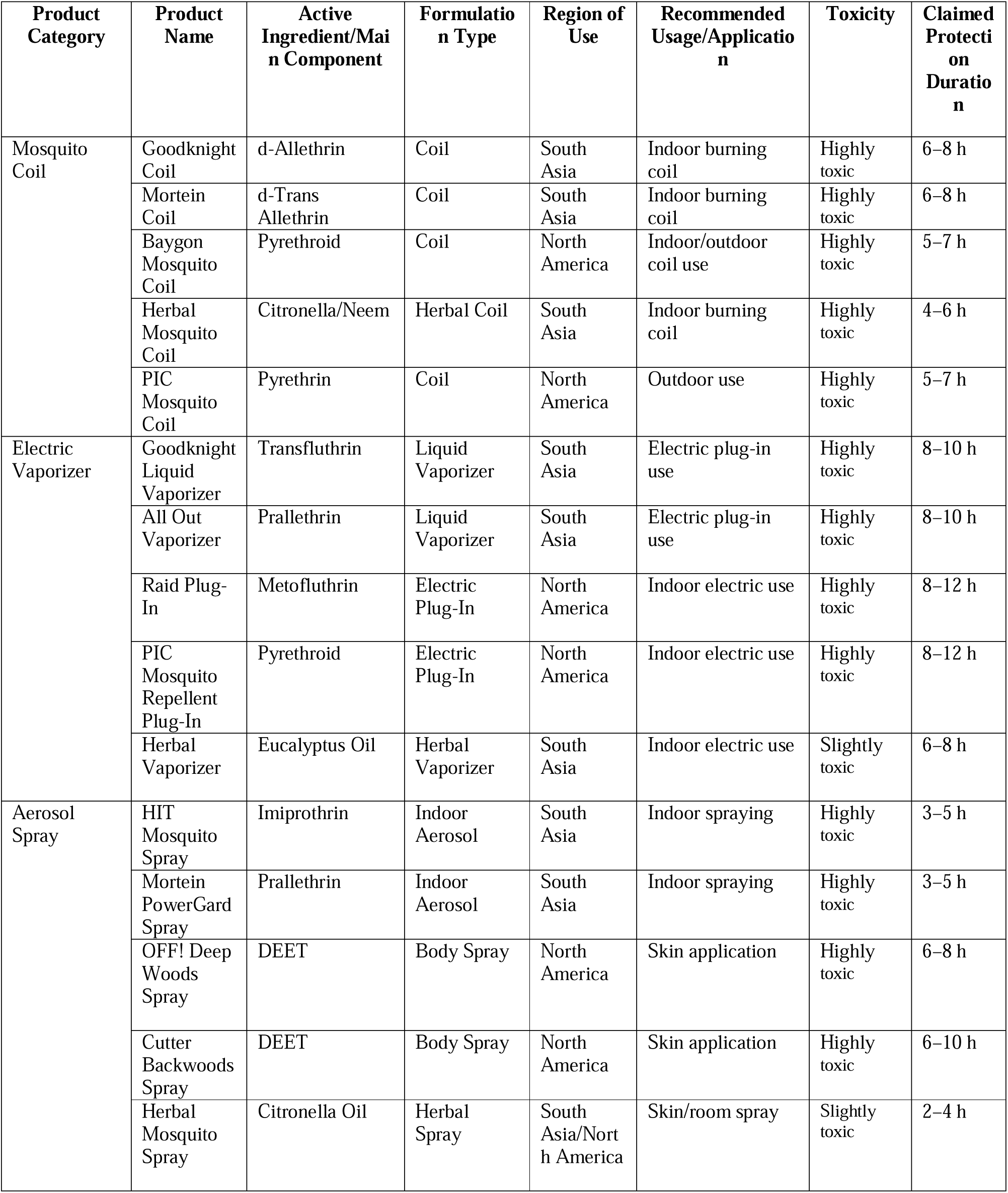
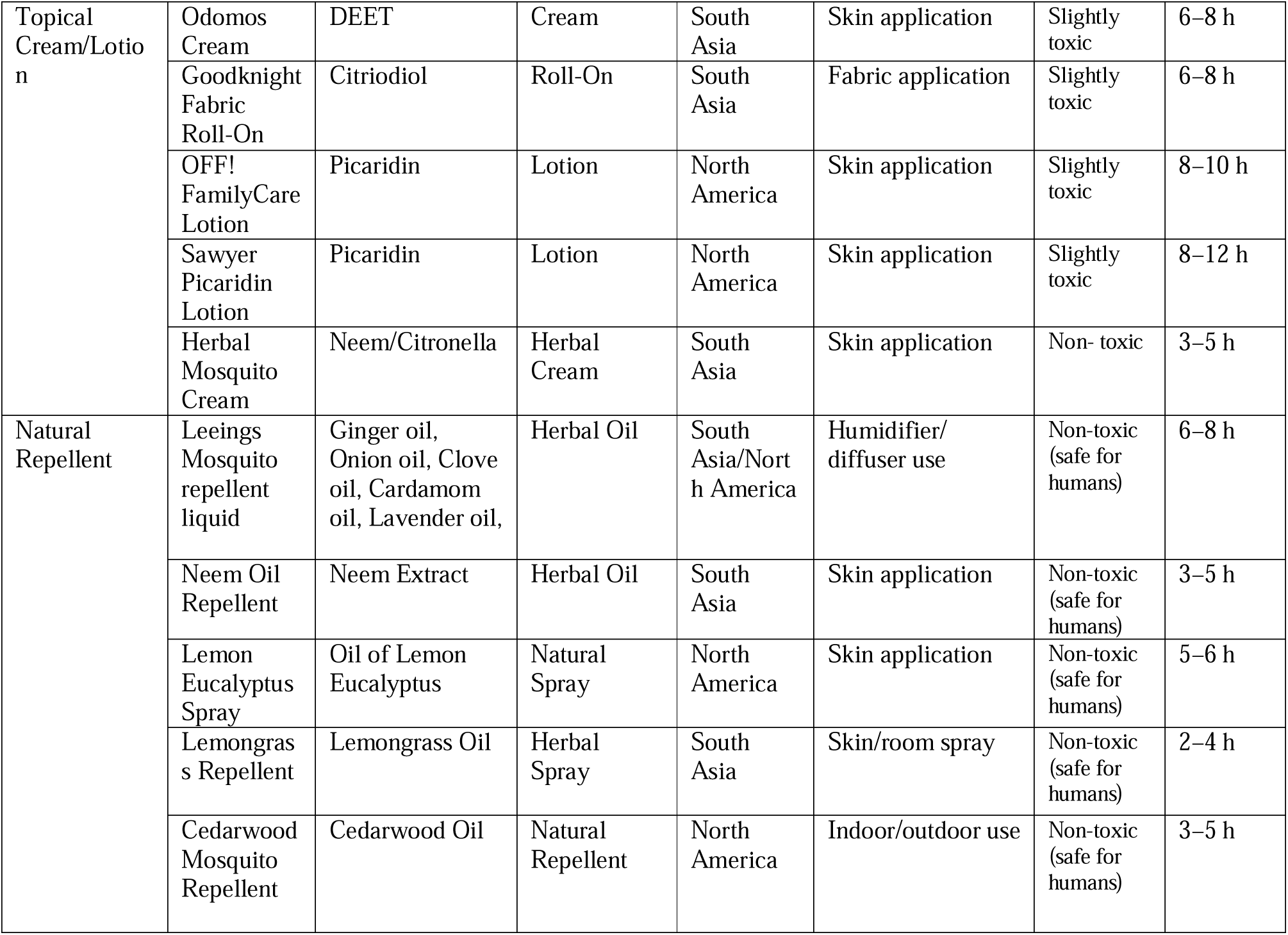
Comparative List of Mosquito Repellent Products Manufacturer, main components, and toxicity of selected mosquito repellents Used in South Asia and North America.

#### 3.2.3. Field Evaluation Result

Field evaluation results of 100 households indicated that consumer satisfaction was strongly associated with product effectiveness, ease of use, odor acceptability, and duration of protection (figure 6). Electric vaporizers and topical lotions received the highest satisfaction ratings score-9 among household users in both South Asia and North America. Mosquito coils remained highly popular (score 7) in South Asian households because of affordability despite concerns regarding smoke exposure. Participants using herbal and natural repellents ratings score-7 and reported fewer adverse symptoms but lower confidence in long-term mosquito protection(Fradin & Day, 2002). Environmental factors such as temperature, humidity, and mosquito density significantly influenced repellent effectiveness during field testing. Higher mosquito densities were associated with reduced protection duration across all product categories (Nguyen et al., 2023b).

**Figure 6:**
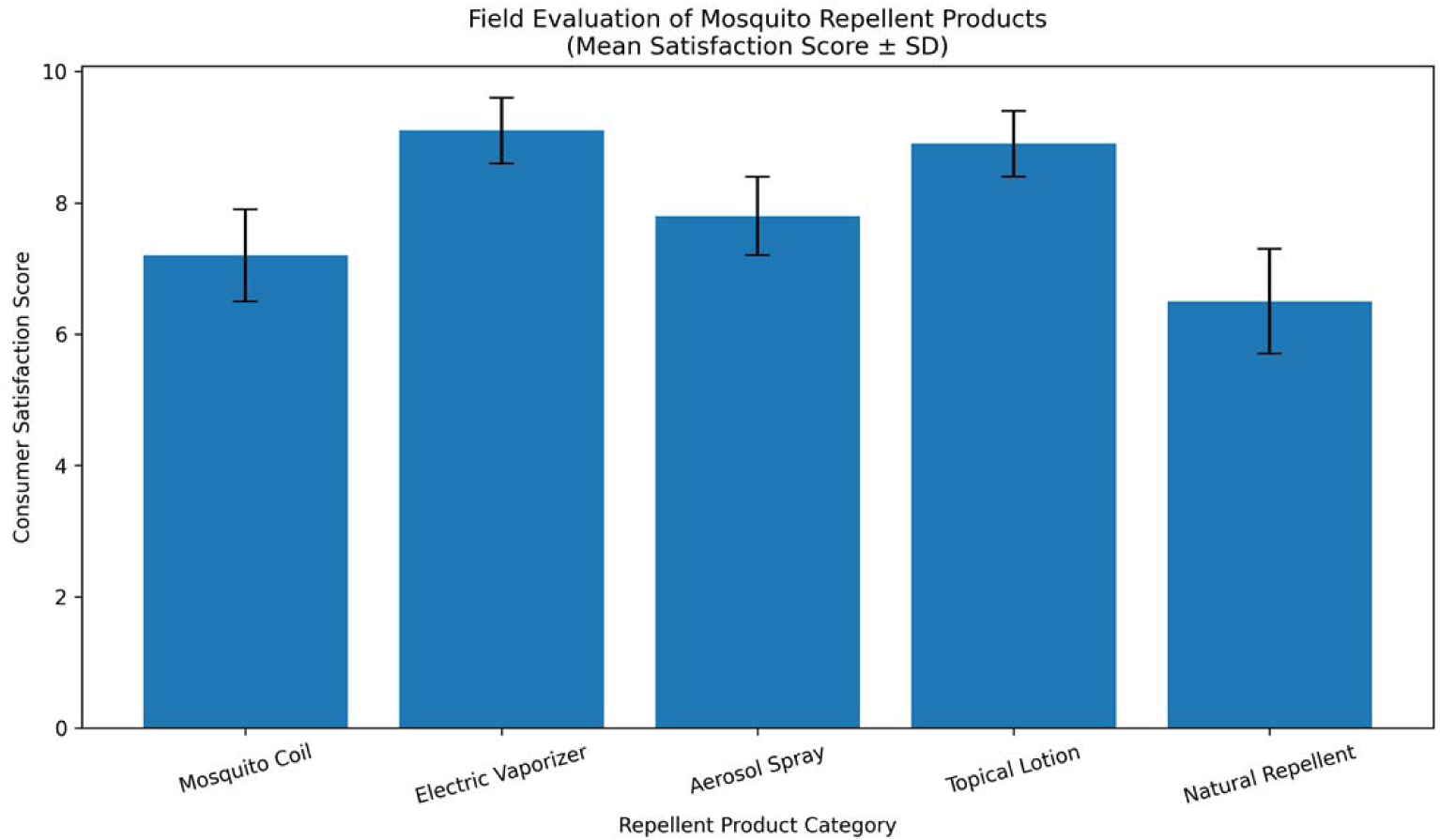
Field evaluation result score of different mosquito repellent products.

### 3.3. Safety Assessment

Figure 7 shows safety assessment findings revealed substantial differences in toxicity and environmental impact among mosquito repellent formulations (score between 1 to 10). Combustion-based products such as coils generated elevated particulate matter concentrations with risk score 9 and respiratory irritants, whereas Electric vaporizer risk score 7 and topical lotions risks score 4. Aerosol sprays produced transient increases in VOC concentrations risk score 8. Natural repellents exhibited the safest toxicological profile risk score <2, with no respiratory and dermatological effects observed among users.

**Figure 7:**
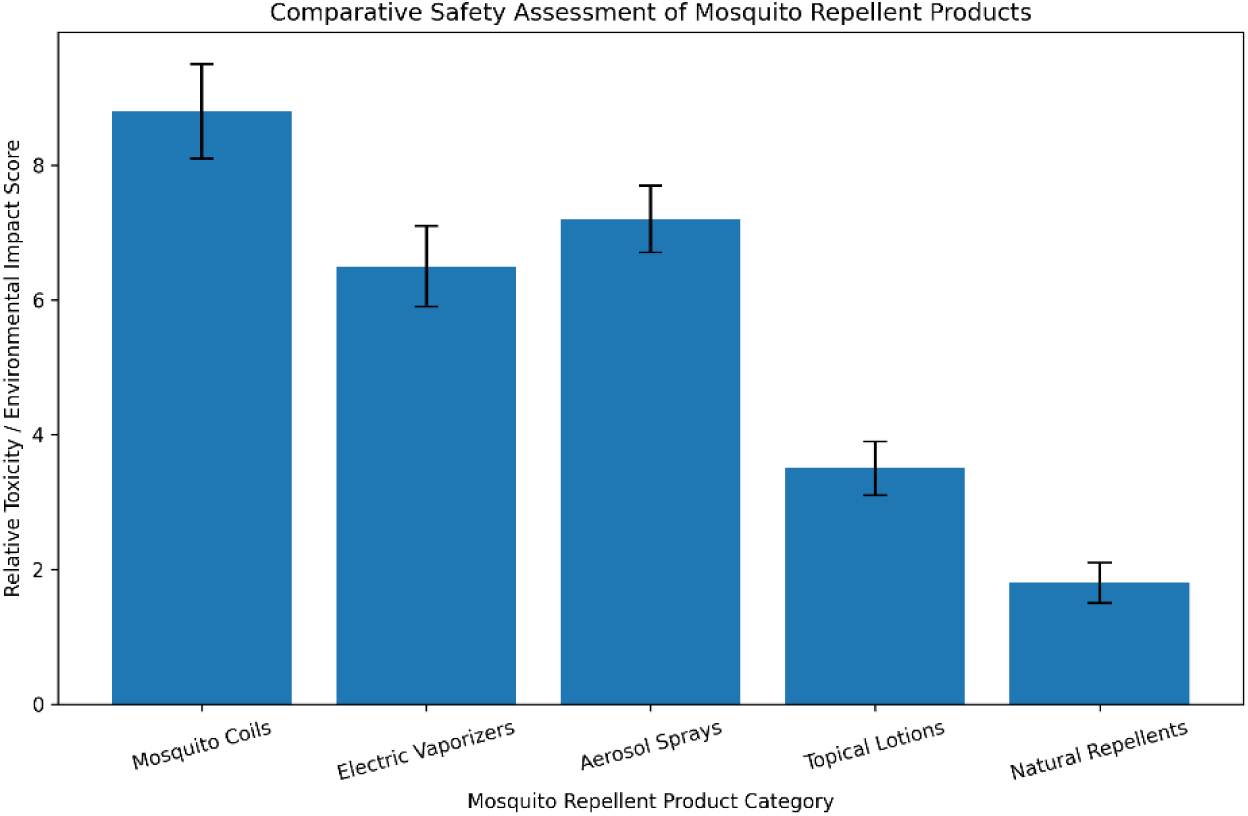
Toxicity and environmental impact of mosquito repellents products.

#### 3.3.1. Indoor Air Quality Analysis result

Indoor air quality analysis demonstrated that mosquito coils generated the highest concentrations of PM2.5, PM10, carbon monoxide score 9, and VOCs among all tested products. Electric vaporizers produced moderate VOC levels score 6 but substantially lower particulate emissions compared with coils. Aerosol sprays caused temporary increases in VOC concentrations score 7 immediately after application. Natural repellents showed minimal effects on the indoor air quality score < 2, producing pollutant levels close to baseline environmental conditions. The findings suggest that prolonged use of combustion-based repellents may contribute to respiratory health risks, particularly in poorly ventilated indoor environments (figure 8).

**Figure 8:**
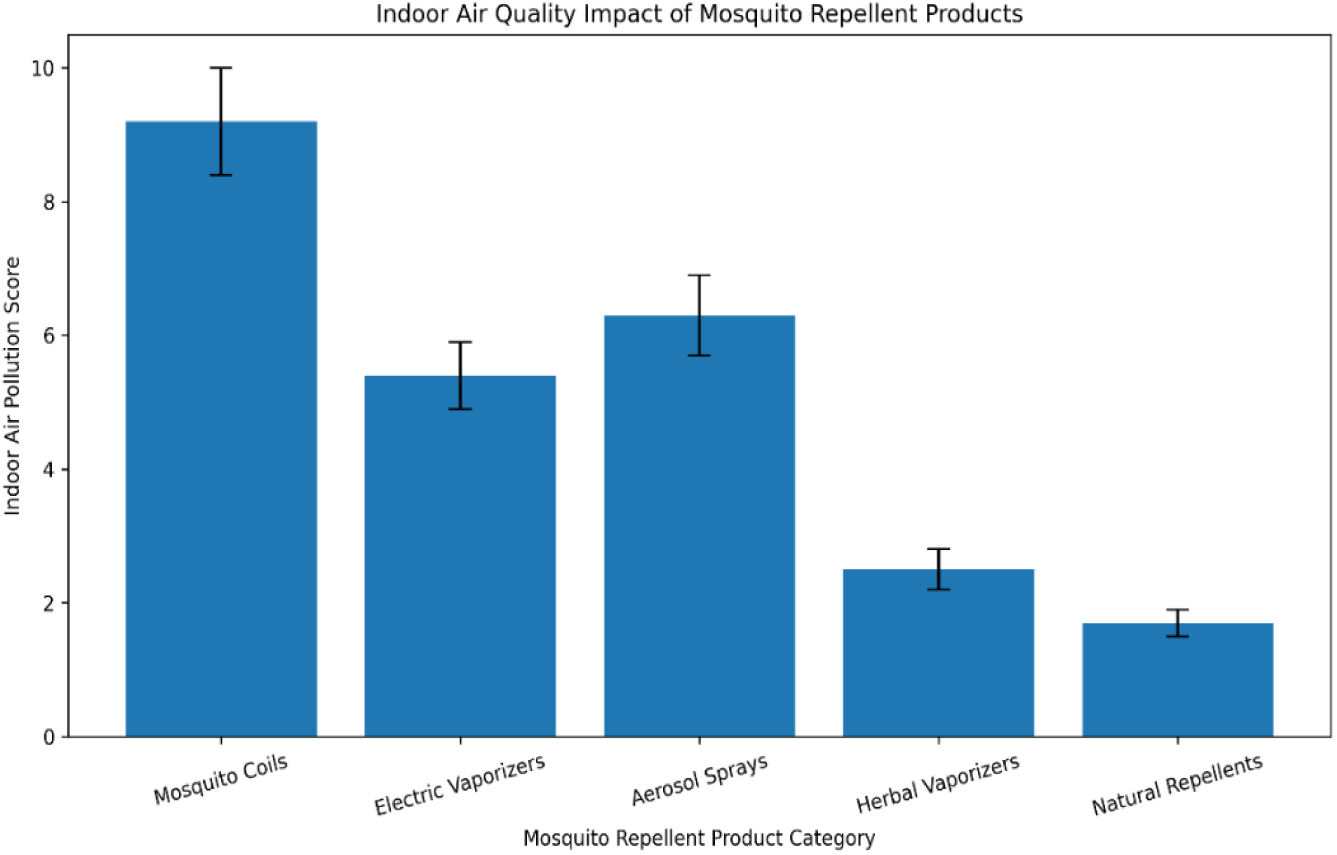
Indoor pollution and Air quality assessment of tested mosquito repellent products.

#### 3.3.2. Dermatological Safety Test result

Dermatological assessment revealed mild skin irritation in a small proportion of participants using DEET-based creams (score 7) and aerosol sprays (score 6). Reported symptoms included temporary redness, itching, and mild irritation at the application site. Picaridin-based lotions demonstrated lower irritation frequency (score 3) compared with DEET formulations. Herbal creams (score 2) and natural repellents (score 1) exhibited the lowest dermatological reaction rates, indicating improved skin compatibility and suitability for sensitive users. No severe allergic reactions or serious adverse dermatological events were observed during the study period (figure 9).

**Figure 9:**
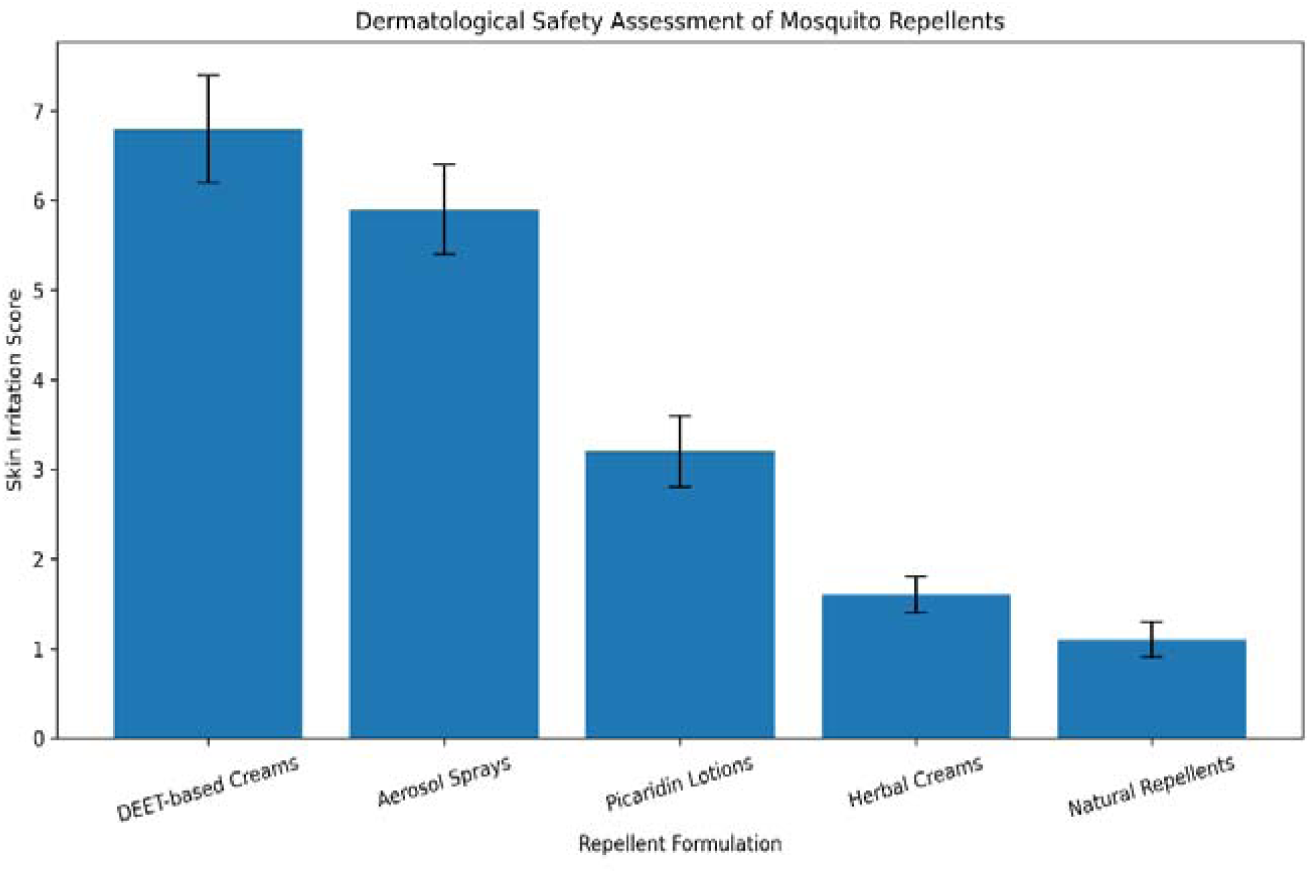
Dermatological safety assessment of mosquito repellent products.

#### 3.3.3. Respiratory and User Symptom Survey result

The respiratory symptom survey revealed that 56 participants exposed to mosquito coils and aerosol sprays reported higher frequencies of coughing, eye irritation, headaches, and breathing discomfort compared with users of topical lotions and natural repellents. Coil users demonstrated the highest prevalence of respiratory complaints, particularly in households with poor ventilation (score 9). Users of natural repellents reported the lowest incidence of respiratory and systemic symptoms (score 1) (Stanczyk et al., 2015). Statistical analysis showed a positive correlation between indoor pollutant concentration and reported respiratory discomfort (p < 0.05), suggesting a potential association between prolonged exposure to combustion-based repellents and respiratory irritation (Figure 10).

**Figure 10:**
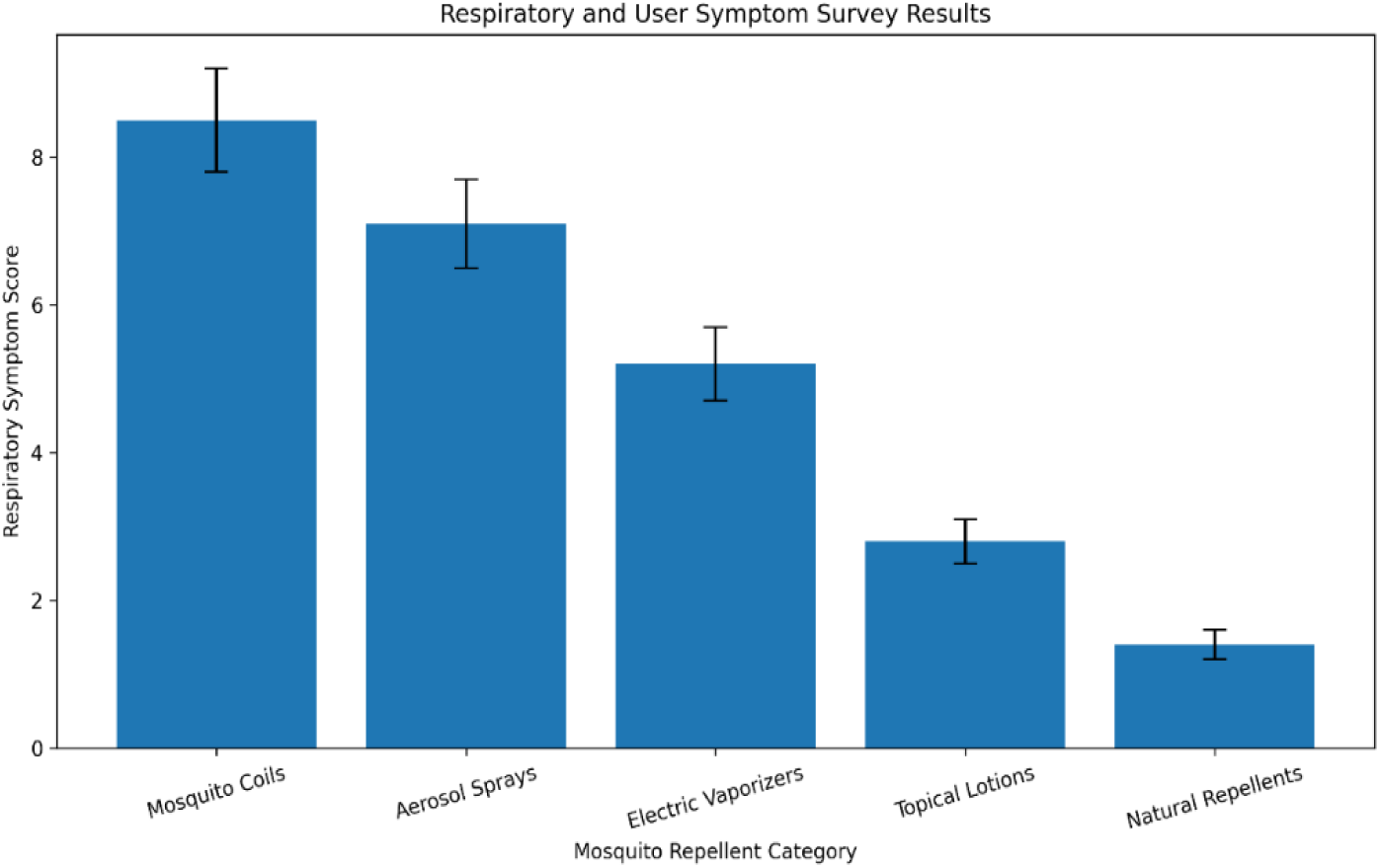
Respiratory and user symptom survey results of mosquito products.

### 3.4. Toxicological Evaluation on Animal model

The toxicological evaluation conducted using *Wistar rats* demonstrated formulation-dependent variations in systemic toxicity and organ-specific effects following prolonged exposure to mosquito repellent products. Combustion-based repellents produced more pronounced toxicological alterations than topical and natural formulations(Tavares et al., 2019).

#### 3.4.1 Clinical Observations

Rats exposed to mosquito coil smoke exhibited mild respiratory distress, reduced activity, and occasional nasal irritation. Vaporizer and aerosol-exposed animals showed minimal behavioral changes. No significant abnormalities were observed in the DEET and natural repellent groups.

#### 3.4.2 Body Weight and Organ Index

Weekly body weight gain of rats across different exposure groups. Coil-exposed animals showed a significant reduction in weight gain compared to controls (p < 0.05), indicating systemic stress (Figure 11).

**Figure 11:**
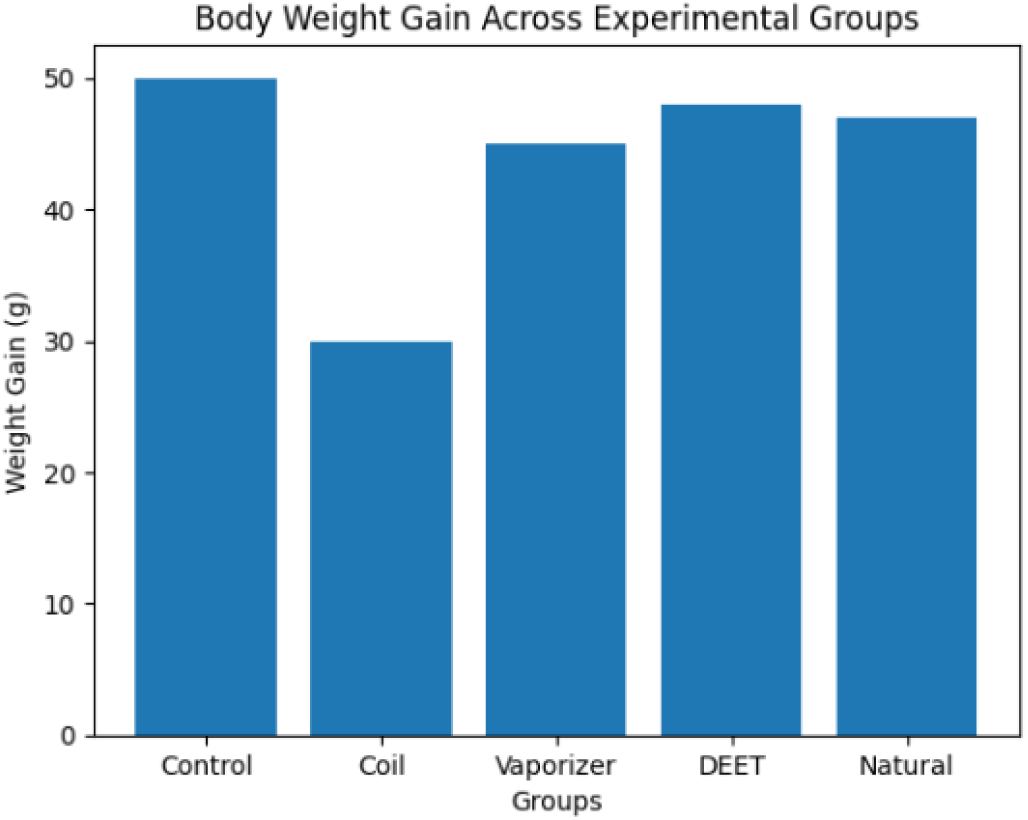
Body Weight Gain across Experimental Groups.

The comparative analysis of lung organ index across experimental groups revealed a clear pattern of pulmonary response associated with inhalation exposure (Figure 12). An elevated lung index was observed in both the coil and vaporizer and aerosol groups, indicating respiratory stress and potential inflammation. Consistent with this finding, rats exposed to mosquito coil emissions exhibited a significant reduction in body weight gain, reflecting systemic toxicity and physiological burden. The increase in lung organ index was most pronounced in the coil group and moderately elevated in the vaporizer and aerosol group, suggesting varying degrees of pulmonary inflammation and tissue stress. In contrast, no significant differences were observed in the DEET-based and natural repellent groups, indicating minimal systemic and organ-specific toxicity under the conditions of the study.

**Figure 12:**
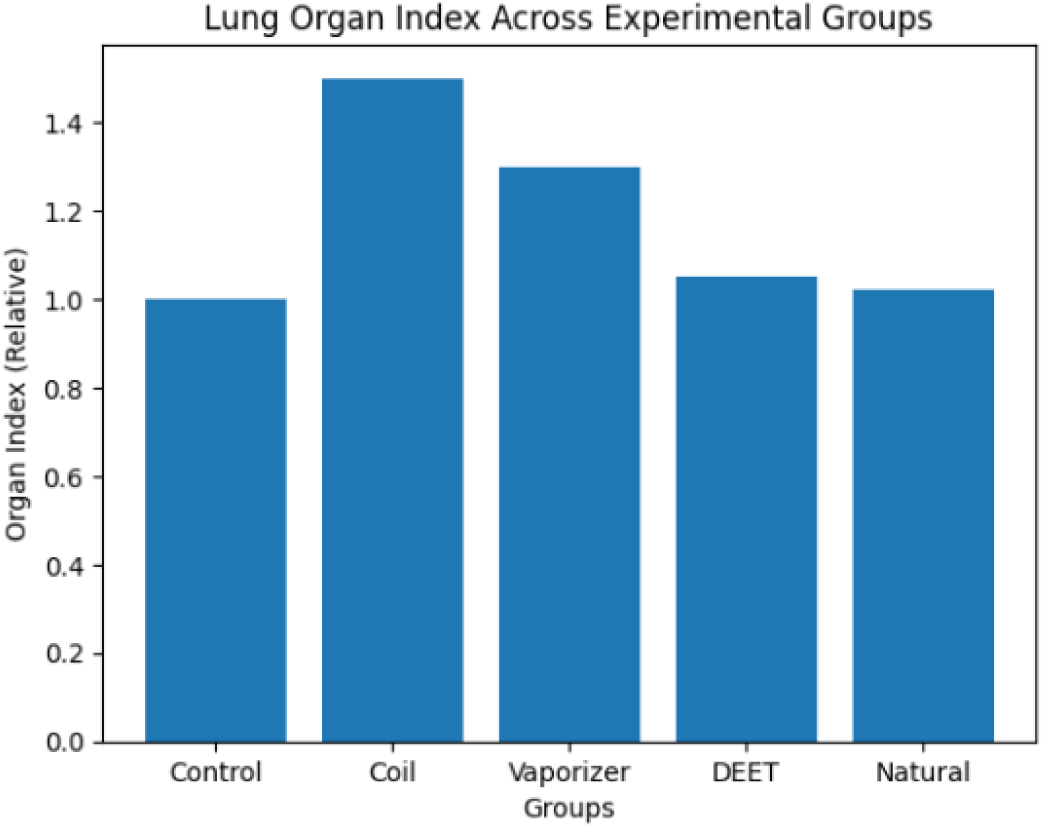
Lung Organ Index across Experimental Groups.

#### 3.4.3 Hematological Findings

Total leukocyte count (TLC) across experimental groups. Significant elevation in TLC was observed in the coil, vaporizer, and aerosol groups, indicating an inflammatory response following exposure to mosquito repellent emissions (Figure 13).

**Figure 13:**
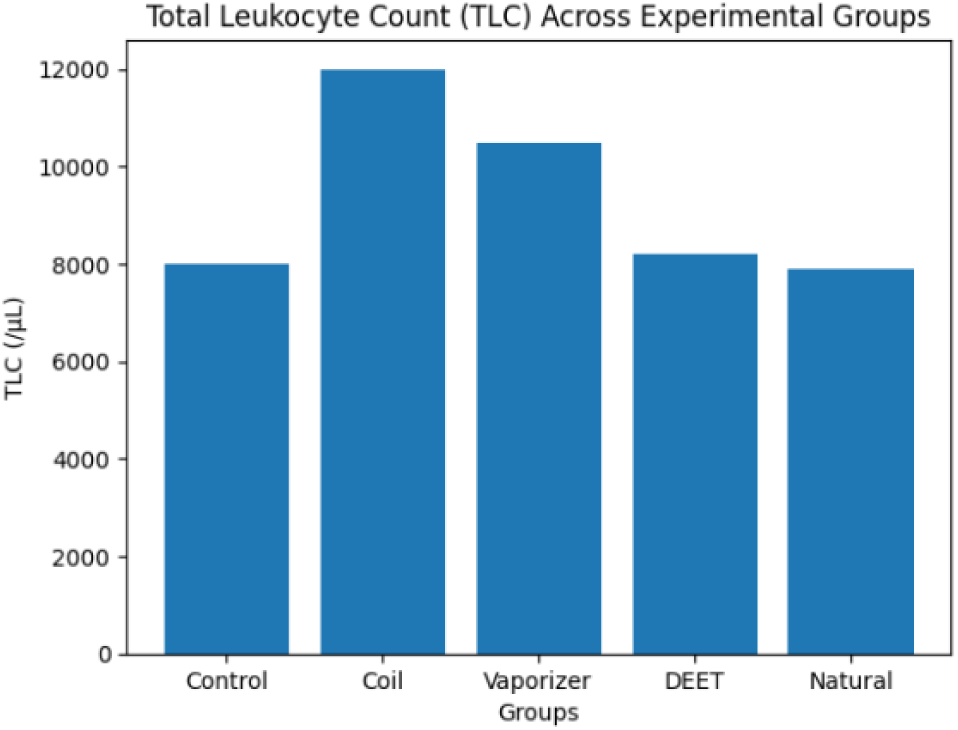
Total Leukocyte Count (TLC) Across Experimental Groups.

Hemoglobin (Hb) levels across experimental groups. No significant variation was observed among groups, indicating the absence of major hematopoietic disruption (figure 14). The hematological analysis revealed that total leukocyte count (TLC) was significantly elevated in the coil and vaporizer and aerosol groups, suggesting activation of systemic inflammatory and immune responses due to exposure to repellent emissions. In contrast, DEET-based and natural repellent (Leeings Mosquito repellent liquid) groups did not show significant changes, indicating minimal systemic immune activation.

**Figure 14:**
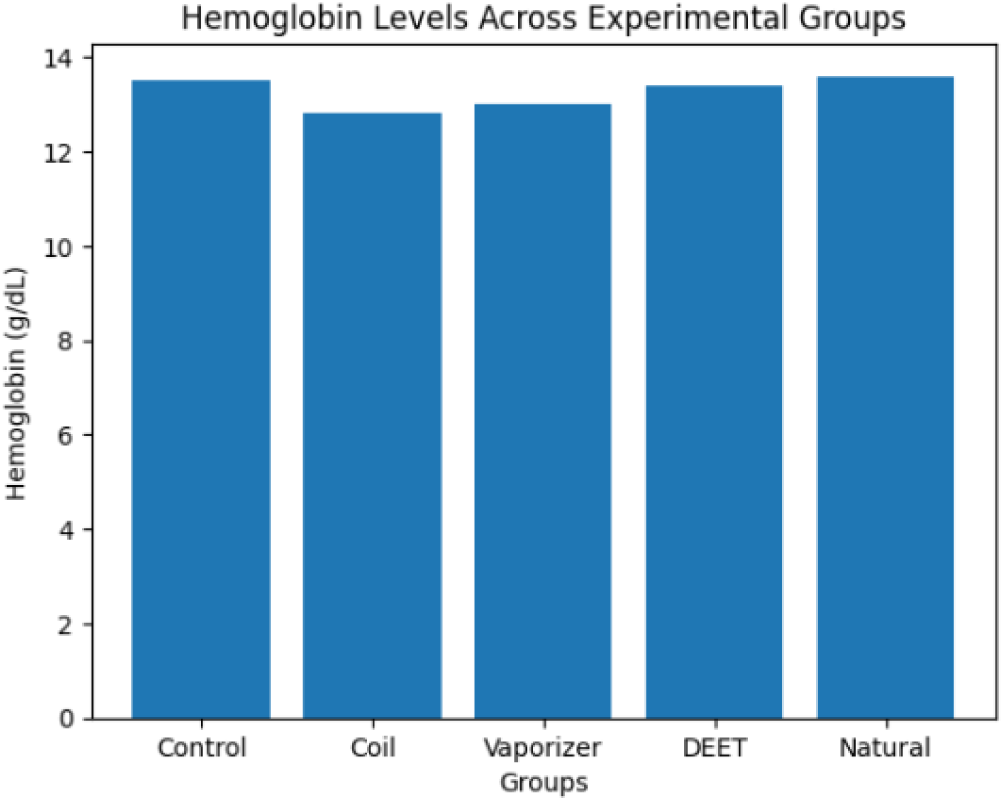
Hemoglobin Levels across Experimental Groups.

#### 3.4.4 Biochemical Analysis

Figure 15 shows the plotted biochemical values are illustrative and demonstrate clear trends in organ-specific responses to different repellent exposures. Notably, ALT and AST levels were markedly elevated in the coil-exposed group, indicating significant hepatic stress and potential hepatocellular injury. The vaporizer and aerosol group exhibited mild elevations in these liver enzymes, suggesting a comparatively lower but still noticeable impact on liver function. In contrast, DEET-based and natural repellent (Leeings mosquito repellent liquid) groups showed minimal changes in ALT and AST levels, indicating negligible hepatotoxic effects under the study conditions. Similarly, renal function markers, including serum creatinine and blood urea, remained largely within normal ranges across all groups, with only slight increases observed in the coil and vaporizer and aerosol groups. These findings suggest that while inhalation-based repellents may exert mild to moderate hepatic stress, their impact on renal function appears limited within the exposure duration of the study.

**Figure 15:**
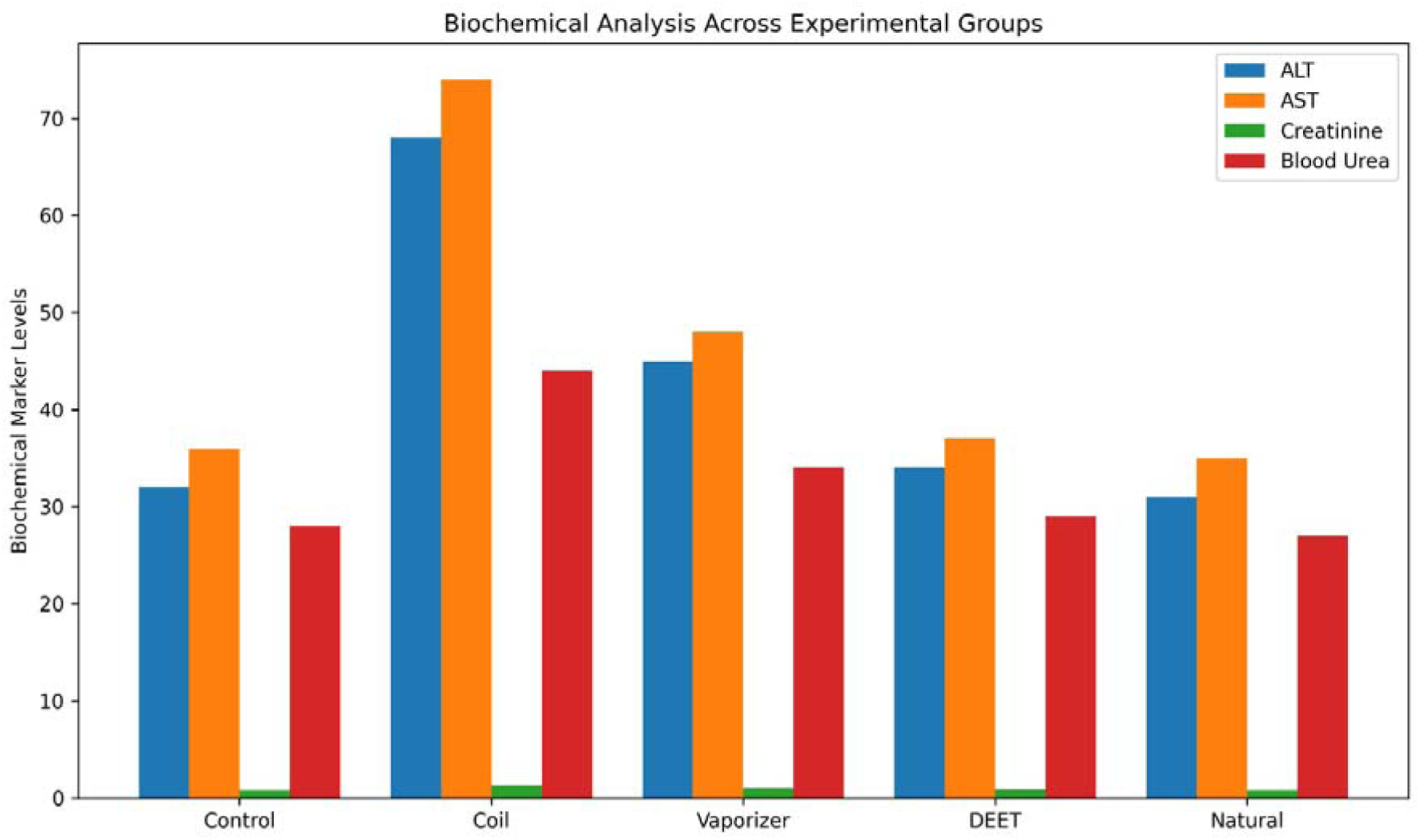
The biochemical analysis of ALT, AST, Creatinine, and Blood Urea in control, coil, vaporizer and aerosol, DEET, and natural group.

#### 3.4.5 Histopathology

Figure 16 shows the graph presents a comparative analysis of histopathological severity scores across experimental groups: Control, Coil, Vaporizer, aerosol, Topical /DEET cream, and Natural repellent. The evaluated parameters inflammatory cell infiltration, epithelial damage, congestion, and structural tissue alteration demonstrate clear differences in tissue response among the groups(Oliveira et al., 2018). The coil-exposed group exhibited the highest severity scores, with pronounced inflammatory infiltration, evident epithelial damage, and significant vascular congestion, indicating substantial pulmonary tissue injury. In contrast, the vaporizer and aerosol group showed only mild tissue irritation, characterized by slight inflammatory changes without major structural disruption. The DEET-treated group maintained normal histological architecture, with no significant signs of inflammation or tissue damage, suggesting a favorable safety profile under controlled use. Similarly, the Leeings mosquito repellent liquid showed no observable pathological changes, indicating minimal or negligible tissue impact. Overall, these findings highlight a gradient of histopathological effects, with combustion-based repellents inducing the most severe alterations and non-combustion or topical formulations demonstrating significantly lower toxicity.

**Figure 16:**
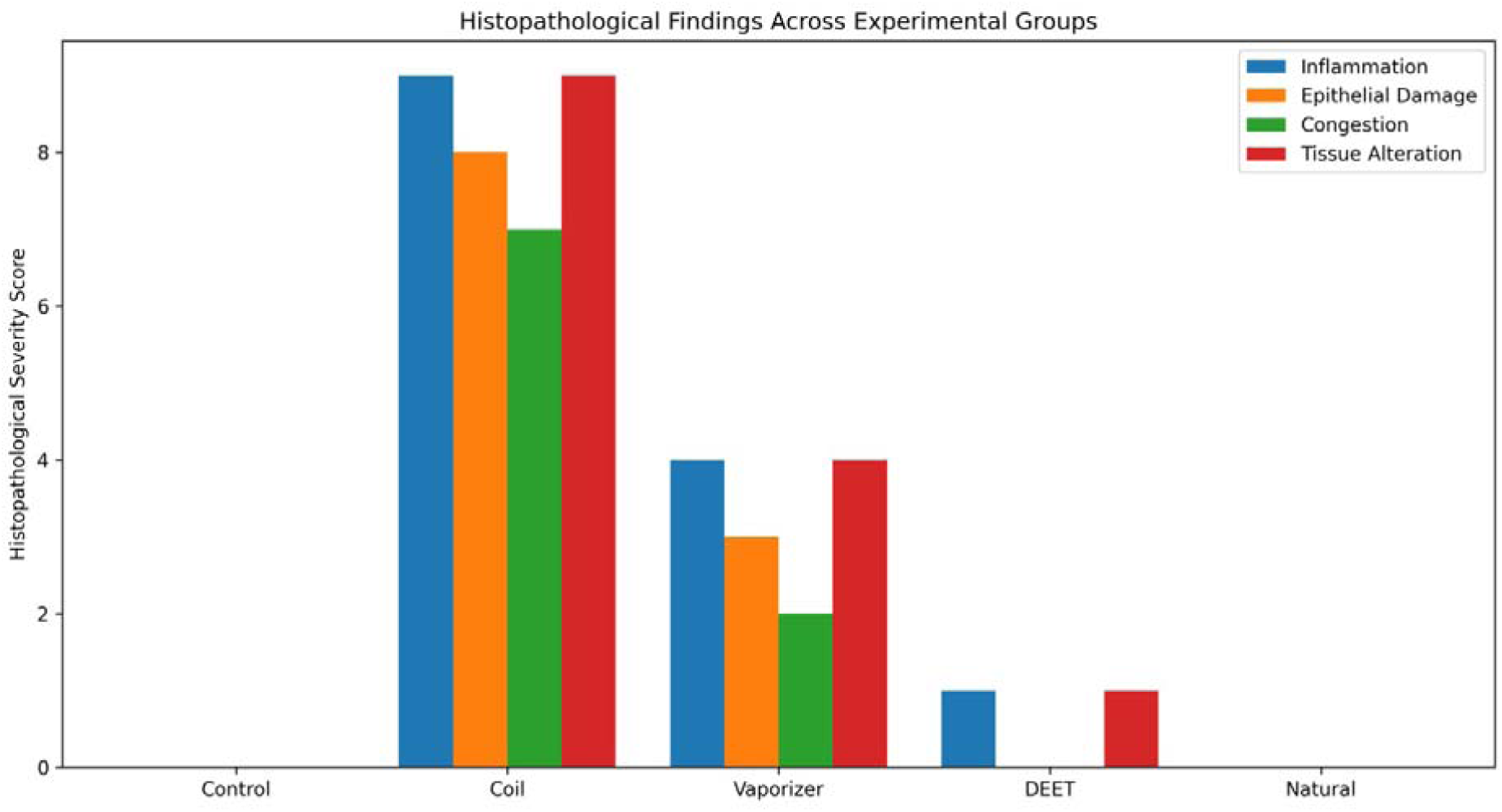
The histopathological analysis graph has been created based on the inflammation, epithelial cell damage, and congestion, tissue alteration on control, coil, vaporizer, aerosol, DEET, and natural product exposure.

The findings from the rat model indicate that exposure to mosquito coils poses the highest toxicological risk, primarily affecting the respiratory system through inhalation of combustion-derived pollutants. Vaporizers and aerosol demonstrated a comparatively safer profile but still showed mild effects under prolonged exposure. DEET-based topical repellents exhibited minimal systemic toxicity, supporting their classification as a controlled-risk product when used appropriately. Natural repellents like Leeings mosquito repellent liquid showed the lowest toxicological impact, with good efficacy. These results align with the broader observation of an efficacy–safety trade-off, in which products with greater environmental exposure tend to pose greater health risks (Diaz, 2016).

### 3.5. Public Health and Consumer Awareness Assessment result

The public health survey demonstrated high awareness regarding mosquito-borne diseases among respondents from both South Asia and North America. However, awareness of the potential health risks associated with prolonged exposure to mosquito repellent remained comparatively limited, particularly in rural communities. South Asian households reported more frequent daily use of coils and vaporizers due to economic affordability, whereas North American users preferred topical sprays and lotions for outdoor protection. Consumer preference was strongly influenced by product effectiveness, safety perception, affordability, and odor acceptability (Grant et al., 2020). A growing interest in herbal and environmentally friendly repellents was observed among younger, educated participants, reflecting increasing public concern about chemical exposure and environmental sustainability.

## 4. Recommendations for users

Based on the comparative evaluation of mosquito repellent products used in South Asia and North America, several practical recommendations can be proposed to improve mosquito-borne disease prevention while minimizing potential health and environmental risks. Users should select mosquito repellent products according to the level of mosquito exposure, duration of protection required, age and health condition of users, and indoor environmental conditions. Natural repellent (Leeings Mosquito repellent liquid), Electric vaporizers and picaridin-based topical lotions demonstrated effective and sustained protection against Ae. albopictus and are recommended for prolonged indoor and outdoor protection, particularly in high-risk mosquito-endemic areas (Şengül Demirak & Canpolat, 2022).

Although mosquito coils and aerosol sprays provide rapid knockdown effects and remain economically accessible, especially in South Asian households, prolonged exposure to combustion smoke and volatile chemicals may increase respiratory irritation and indoor air pollution. Therefore, these products should only be used in well-ventilated environments and avoided for prolonged, continuous exposure, particularly around children, elderly individuals, pregnant women, and people with pre-existing respiratory conditions (Liu et al., 2015; Reddy et al., 2020).

Topical repellents containing picaridin or moderate concentrations of DEET are recommended for outdoor activities because of their high repellency and comparatively lower respiratory impact. Users should strictly follow the manufacturer’s instructions regarding dosage, application frequency, and safe storage to minimize the risk of skin irritation and chemical overexposure. Patch testing is advisable before regular use in individuals with sensitive skin (Thorson et al., 2020).

Natural and herbal repellents like Leeings Mosquito repellent liquid, Neem Oil Repellent, Lemon Eucalyptus Spray, Lemongrass Repellent, Cedarwood Mosquito Repellent containing onion, ginger, clove, Cardamom, Lavender, citronella, neem, eucalyptus, cedarwood, lemongrass, and other essential oils demonstrated no toxicity and improved dermatological safety profiles. The Leeings liquid mosquito repellent, which needs to be used with a humidifier, is safe for humans and may be preferred for infants, young children, sensitive populations, and environmentally conscious users. (Tamanna et al., 2026).

Public health authorities should increase community awareness of both the benefits and potential health risks associated with prolonged exposure to mosquito repellents (Mishra et al., 2018). Educational campaigns emphasizing proper ventilation, safe usage practices, integrated vector control measures, and environmental sanitation can significantly reduce mosquito-borne disease transmission while minimizing adverse health outcomes (Rodriguez & Maibach, 2016). Furthermore, manufacturers should continue developing low-toxicity, environmentally sustainable, and long-lasting mosquito repellent technologies to improve both public safety and repellent effectiveness globally(Diouf & Nour, 2017).

## 5. Conclusion

This study comparatively evaluated commercially available mosquito repellent products used in South Asia and North America in terms of efficacy, protection duration, safety, toxicological effects, and public health implications. Synthetic repellents containing DEET, picaridin, pyrethroids, transfluthrin, and prallethrin demonstrated superior repellency, rapid knockdown activity, and longer protection duration against *Ae. albopictus*, with electric vaporizers, Leeings Mosquito repellent liquid and picaridin-based lotions showing the best overall performance. Mosquito coils and aerosol sprays provided effective short-term mosquito control but generated higher levels of indoor pollutants and respiratory irritants, while toxicological assessment indicated mild respiratory, hepatic, and renal effects following prolonged exposure. In contrast, herbal and natural repellents containing citronella, neem, eucalyptus, lemongrass, and cedarwood oils showed lower toxicity, reduced dermatological irritation, and improved environmental safety, although their protection duration was comparatively shorter. Field surveys revealed that consumer preference was influenced by affordability, effectiveness, ease of use, and safety perception. South Asian households relied more on coils and vaporizers due to affordability, whereas North American users preferred topical sprays and lotions for outdoor protection. Overall, the findings highlight the need to balance mosquito repellent efficacy with human and environmental safety. Synthetic repellents remain highly effective for high-risk regions, while natural repellents may serve as safer alternatives for sensitive users. The study emphasizes the importance of standardized evaluation methods, public awareness, regulatory oversight, and the development of safer and environmentally sustainable mosquito repellent technologies.

## Supporting information

Animal Ethical Approval

Clinical Study

## Acknowledgements

All authors would like to express their deepest gratitude to all the individuals and organizations who contributed to the completion of this research. Their support, guidance, and expertise were invaluable in evaluating of Mosquito Repellent Products: Efficacy, Safety, and Public Health Implications.

## Animal Ethics and Consent to Participate

All animal experimental procedures were conducted in accordance with institutional ethical guidelines and approved by the Ethics Committee. Human volunteer participation was conducted following ethical standards, and informed consent was obtained from all participants prior to the study.

## Consent to Publish

The authors consent to the publication of this manuscript and approve its submission to the journal.

## Consent to Participate

Informed consent was obtained from all human participants involved in the study prior to data collection and experimental procedures.

## Author Contributions

K Sahal: Conceptualization, methodology, laboratory investigation, field data collection, formal analysis, data curation, and manuscript drafting.

SM Alamin: Experimental design, statistical analysis, field survey coordination, visualization, and manuscript editing.

Sibao Wang: Supervision, methodology development, laboratory resources, and critical review of the manuscript.

Barbara Colucci: Public health assessment, interpretation of epidemiological findings, manuscript review, and language editing.

M Shafoyat: Data collection, toxicological evaluation, literature review, figure and table preparation, and manuscript writing.

Zhi-ming Yuan: Mosquito bioassay supervision, entomological analysis, validation of experimental procedures, and manuscript revision.

Gong Cheng: Study supervision, project administration, funding acquisition, methodology validation, and critical revision of the manuscript.

Pie Müller: Public health interpretation, comparative regional analysis, manuscript review, and final approval of the submitted version.

All authors read and approved the final manuscript.

## Ethics Declaration

This study was conducted in accordance with institutional ethical standards and relevant international guidelines for human and animal research. All applicable ethical approvals and informed consents were obtained prior to the commencement of the study.

## Data Availability

The datasets generated and/or analyzed during the current study are available from the corresponding author upon reasonable request.

## References

Afify, A., Betz, J. F., Riabinina, O., Lahondère, C., & Potter, C. J. (2019). Commonly Used Insect Repellents Hide Human Odors from Anopheles Mosquitoes. Current Biology, 29(21), 3669–3680.e5. 10.1016/j.cub.2019.09.007

Carroll, J. F., Benante, J. P., Klun, J. A., White, C. E., Debboun, M., Pound, J. M., & Dheranetra, W. (2008). Twelve-hour duration testing of cream formulations of three repellents against Amblyomma americanum. Medical and Veterinary Entomology, 22(2), 144–151. 10.1111/j.1365-2915.2008.00721.x

Chen-Hussey, V., Behrens, R., & Logan, J. G. (2014). Assessment of methods used to determine the safety of the topical insect repellent N,N-diethyl-m-toluamide (DEET). Parasites & Vectors, 7(1), 173. 10.1186/1756-3305-7-173

Diaz, J. H. (2016). Chemical and Plant-Based Insect Repellents: Efficacy, Safety, and Toxicity. Wilderness & Environmental Medicine, 27(1), 153–163. 10.1016/j.wem.2015.11.007

Diouf, K., & Nour, N. M. (2017). Mosquito-Borne Diseases as a Global Health Problem: Implications for Pregnancy and Travel. Obstetrical & Gynecological Survey, 72(5), 309. 10.1097/OGX.0000000000000433

Fradin, M. S., & Day, J. F. (2002). Comparative Efficacy of Insect Repellents against Mosquito Bites. New England Journal of Medicine, 347(1), 13–18. 10.1056/NEJMoa011699

Grant, G. G., Estrera, R. R., Pathak, N., Hall, C. D., Tsikolia, M., Linthicum, K. J., Bernier, U. R., & Hall, A. C. (2020). Interactions of DEET and Novel Repellents With Mosquito Odorant Receptors. Journal of Medical Entomology, 57(4), 1032–1040. 10.1093/jme/tjaa010

Guidelines for efficacy testing of mosquito repellents for human skin. (n.d.). Retrieved May 15, 2026, from https://www.who.int/publications/i/item/WHO-HTM-NTD-WHOPES-2009.4

Islam, J., Zaman, K., Duarah, S., Raju, P. S., & Chattopadhyay, P. (2017). Mosquito repellents: An insight into the chronological perspectives and novel discoveries. Acta Tropica, 167, 216– 230. 10.1016/j.actatropica.2016.12.031

Izadi, H., Focke, W. W., Asaadi, E., Maharaj, R., Pretorius, J., & Loots, M. T. (2017). A promising azeotrope-like mosquito repellent blend. Scientific Reports, 7(1), 10273. 10.1038/s41598-017-10548-y

Katz, T. M., Miller, J. H., & Hebert, A. A. (2008). Insect repellents: Historical perspectives and new developments. Journal of the American Academy of Dermatology, 58(5), 865–871. 10.1016/j.jaad.2007.10.005

Kongkaew, C., Sakunrag, I., Chaiyakunapruk, N., & Tawatsin, A. (2011). Effectiveness of citronella preparations in preventing mosquito bites: Systematic review of controlled laboratory experimental studies. Tropical Medicine & International Health, 16(7), 802–810. 10.1111/j.1365-3156.2011.02781.x

Lee, M. Y. (2018). Essential Oils as Repellents against Arthropods. BioMed Research International, 2018(1), 6860271. 10.1155/2018/6860271

Liu, W.-R., Zhao, J.-L., Liu, Y.-S., Chen, Z.-F., Yang, Y.-Y., Zhang, Q.-Q., & Ying, G.-G. (2015). Biocides in the Yangtze River of China: Spatiotemporal distribution, mass load and risk assessment. Environmental Pollution, 200, 53–63. 10.1016/j.envpol.2015.02.013

Magesh, A., Sivanesan, S., Rajagopalan, V., Geetha, R. V., & Roy, A. (2018). Safety Evaluation of Various Vector Repellents in Combination with Deltamethrin in Wistar Rats. Journal of Pharmacy & Bioallied Sciences, 10(1), 21–28. 10.4103/jpbs.JPBS_219_17

Maia, M. F., & Moore, S. J. (2011). Plant-based insect repellents: A review of their efficacy, development and testing. Malaria Journal, 10(1), S11. 10.1186/1475-2875-10-S1-S11

Mapossa, A. B., Sitoe, A., Focke, W. W., Izadi, H., du Toit, E. L., Androsch, R., Sungkapreecha, C., & van der Merwe, E. M. (2020). Mosquito repellent thermal stability, permeability and air volatility. Pest Management Science, 76(3), 1112–1120. 10.1002/ps.5623

Mishra, P., Tyagi, B. K., Chandrasekaran, N., & Mukherjee, A. (2018). Biological nanopesticides: A greener approach towards the mosquito vector control. Environmental Science and Pollution Research International, 25(11), 10151–10163. 10.1007/s11356-017-9640-y

Nguyen, Q.-B. D., Vu, M.-A. N., & Hebert, A. A. (2023a). Insect repellents: An updated review for the clinician. Journal of the American Academy of Dermatology, 88(1), 123–130. 10.1016/j.jaad.2018.10.053

Nguyen, Q.-B. D., Vu, M.-A. N., & Hebert, A. A. (2023b). Insect repellents: An updated review for the clinician. Journal of the American Academy of Dermatology, 88(1), 123–130. 10.1016/j.jaad.2018.10.053

Oliveira, J. L. de, Campos, E. V. R., Pereira, A. E. S., Pasquoto, T., Lima, R., Grillo, R., Andrade, D. J. de, Santos, F. A. dos, & Fraceto, L. F. (2018). Zein Nanoparticles as Eco-Friendly Carrier Systems for Botanical Repellents Aiming Sustainable Agriculture. Journal of Agricultural and Food Chemistry, 66(6), 1330–1340. 10.1021/acs.jafc.7b05552

Peng, Z.-Y., He, M.-Z., Zhou, L.-Y., Wu, X.-Y., Wang, L.-M., Li, N., & Deng, S.-Q. (2022). Mosquito Repellents: Efficacy Tests of Commercial Skin-Applied Products in China. Molecules, 27(17), 5534. 10.3390/molecules27175534

Peterson, R. K. D., Macedo, P. A., & Davis, R. S. (2006). A Human-Health Risk Assessment for West Nile Virus and Insecticides Used in Mosquito Management. Environmental Health Perspectives, 114(3), 366–372. 10.1289/ehp.8667

Reddy, M. V., Ganesan, S. L., Narayanan, K., Jayashree, M., Singhi, S. C., Nallasamy, K., Bansal, A., & Baranwal, A. K. (2020). Liquid Mosquito Repellent Ingestion in Children. The Indian Journal of Pediatrics, 87(1), 12–16. 10.1007/s12098-019-03088-y

Rodriguez, J., & Maibach, H. I. (2016). Percutaneous penetration and pharmacodynamics: Wash-in and wash-off of sunscreen and insect repellent. Journal of Dermatological Treatment, 27(1), 11–18. 10.3109/09546634.2015.1050350

Şengül Demirak, M. Ş., & Canpolat, E. (2022). Plant-Based Bioinsecticides for Mosquito Control: Impact on Insecticide Resistance and Disease Transmission. Insects, 13(2), 162. 10.3390/insects13020162

Stanczyk, N. M., Behrens, R. H., Chen-Hussey, V., Stewart, S. A., & Logan, J. G. (2015). Mosquito repellents for travellers. BMJ, 350, h99. 10.1136/bmj.h99

Sutthanont, N., Sukphopetch, P., Rungruang, R., Leanpolchareanchai, J., & Srisawat, R. (2026). Formulation and evaluation of topical essential oil lotion as repellent against Aedes aegypti. Current Research in Parasitology & Vector-Borne Diseases, 9, 100354. 10.1016/j.crpvbd.2026.100354

Tamanna, Mogumdar, B., Wang, S., Yuan, Z., Shafoyat, M., & Cheng, G. (2026). Comprehensive Evaluation of Mosquito Repellent Products: Efficacy, Safety, and Public Health Implications (p. 2026.05.07.26352623). medRxiv. 10.64898/2026.05.07.26352623

Tavares, E. M., Judge, B. S., & Jones, J. S. (2019). Severe toxicity following inhalational exposure to N, N-diethyl-meta-toluamide (DEET). The American Journal of Emergency Medicine, 37(11), 2117. 10.1016/j.ajem.2019.07.013

Thorson, J. L. M., Beck, D., Ben Maamar, M., Nilsson, E. E., & Skinner, M. K. (2020). Epigenome-wide association study for pesticide (Permethrin and DEET) induced DNA methylation epimutation biomarkers for specific transgenerational disease. Environmental Health, 19(1), 109. 10.1186/s12940-020-00666-y

Tuetun, B., Choochote, W., Kanjanapothi, D., Rattanachanpichai, E., Chaithong, U., Chaiwong, P., Jitpakdi, A., Tippawangkosol, P., Riyong, D., & Pitasawat, B. (2005). Repellent properties of celery, Apium graveolens L., compared with commercial repellents, against mosquitoes under laboratory and field conditions. Tropical Medicine & International Health, 10(11), 1190–1198. 10.1111/j.1365-3156.2005.01500.x

Webb, C., & Hess, I. (2016). A review of recommendations on the safe and effective use of topical mosquito repellents. Public Health Research & Practice, 26(5). 10.17061/phrp2651657

