## Supplementary material for "Comparative Evaluation of Mosquito Repellent Products in South Asia and North America: Efficacy, Safety, and Public Health Implications": Supplymentry materials: Ethical approval Animal file.pdf

### ETHICAL APPROVAL APPLICATION

**Animal Research (Rat Model)**

#### 1. Title of the Study

**Safety and Toxicological Evaluation of Mosquito Repellent Products in a Rat Model**

#### 2. Principal Investigator (PI)


- Name: Md Shafoyat
- Designation: Professor
- Department: Biomedical Enigneering
- Institution: Military Institute of Science and Technology
-


#### 3. Project Duration

Start Date: 01 Jun 2025
End Date: 01 Jan 2026
Total Duration: 06 months

#### 4. Background and Rationale

Mosquito repellent products are widely used for protection against vector-borne diseases such as dengue, malaria, and chikungunya. Despite their widespread use, limited studies have evaluated their systemic toxicity and safety profiles under controlled experimental conditions. This study aims to assess the potential toxicological effects of selected mosquito repellent formulations using a rat model to ensure safety for human exposure and guide public health recommendations.

#### 5. Objectives

**General Objective:**

To evaluate the safety and toxicological effects of mosquito repellent products in a rat model.

**Specific Objectives:**

- To assess acute and sub-acute toxicity of selected mosquito repellents
- To evaluate hematological and biochemical changes in treated rats
- To observe histopathological changes in vital organs (liver, kidney, lung)
- To compare toxicity profiles between different repellent formulations

#### 6. Methodology

##### 6.1 Study Design

Experimental laboratory-based animal study.

##### 6.2 Animal Model

- Species: Albino Wistar rats (Rattus norvegicus)
- Sex: Male/Female (as applicable)
- Weight: 150–250 g
- Number of animals: 15

##### 6.3 Grouping of Animals

Animals were randomly divided into five groups (n = 3 per group):

- **Group I (Control)**
- **Group II (Coil Exposure)**
- **Group III (Vaporizer** and aerosol **Exposure)**
- **Group IV (DEET Application)**
- **Group V (Natural Repellent)**

The exposure duration was maintained for **28 consecutive days** to assess sub-chronic toxicity.

##### 6.4 Test Substances

Mosquito repellent products (specify active ingredients):

- **Group I (Control):** No exposure
- **Group II (Coil Exposure):** Exposure to mosquito coil smoke (8 hours/day)
- **Group III (Vaporizer** and aerosol **Exposure):** Exposure to vaporizer and aerosol emissions (8 hours/day)
- **Group IV (DEET Application):** Topical application of DEET-based cream (standard dermal dose)
- **Group V (Natural Repellent):** Exposure using a humidifier (8 hours/day)

##### 6.5 Route of Exposure

- Dermal application / inhalation exposure (specify clearly)
- Duration: Acute (8 hrs day) and Sub-acute (28 days)

#### 6.6 Outcome Measures

- Body weight changes
- Behavioral observations
- Hematological parameters (Hb, WBC, RBC, etc.)
- Biochemical markers (ALT, AST, creatinine, urea)
- Histopathological examination of organs

#### 6.7 Ethical Considerations, Anesthesia, and Humane Endpoints

All animal experiments were conducted in accordance with institutional ethical guidelines and internationally accepted principles for the care and use of laboratory animals. .

(a) Methods of sacrifice

At the end of the experimental period, animals were humanely sacrificed under deep anesthesia to minimize pain and distress. Euthanasia was performed by overdose administration of ketamine–xylazine followed by cervical dislocation to ensure rapid and irreversible death prior to tissue collection. This method was selected in accordance with standard laboratory animal welfare recommendations.

(b) Methods of anesthesia and/or analgesia, and

For procedures involving handling and sample collection, animals were anesthetized using an intraperitoneal injection of Ketamine (80–100 mg/kg body weight) combined with Xylazine (5–10 mg/kg body weight) to ensure adequate anesthesia, analgesia, and muscle relaxation during experimental procedures. The depth of anesthesia was monitored through assessment of pedal withdrawal and corneal reflexes prior to invasive manipulations.

(c) Efforts to alleviate suffering.

Throughout the study, several measures were implemented to alleviate animal suffering, including daily health monitoring, maintenance of appropriate housing conditions, provision of adequate nutrition and hydration, and minimization of unnecessary handling. Animals exhibiting severe distress, persistent respiratory difficulty, excessive weight loss, or signs of severe pain were designated for immediate humane intervention or euthanasia according to predefined humane endpoint criteria.

#### 6.7 Data Analysis

Statistical analysis will be performed using appropriate software (e.g., SPSS/GraphPad Prism). Results will be expressed as mean ± SD and analyzed using ANOVA or t-tests with significance level $p < 0.05$.

**7. Animal Welfare Considerations**

- Animals will be housed in standard laboratory conditions (temperature, humidity, 12-hour light/dark cycle).
- Standard feed and water will be provided ad libitum.
- All procedures will follow internationally accepted animal welfare guidelines (e.g., ARRIVE guidelines / OECD / institutional animal care policies).
- Minimal pain and distress will be ensured.
- If required, anesthesia or humane euthanasia will be performed using approved protocols.

**8. Expected Benefits**

- Identification of potential toxicity of commonly used mosquito repellents
- Contribution to public health safety guidelines
- Support for regulatory evaluation of repellent formulations

**9. Potential Risks**

- Minimal risk to animals due to chemical exposure
- Proper monitoring and humane handling will reduce suffering

**10. Conflict of Interest**

No conflict of interest

**11. Funding Source**

- Self-funded

**12. Declaration by Principal Investigator**

I hereby declare that the above information is correct and that the study will be conducted according to ethical guidelines for animal research. All necessary precautions will be taken to minimize animal suffering.

Signature: ______________________

Name: Prof Md Shafoyat

Date: 01 Jun 2025

#### 13. Ethical Review Committee Section

- Institutional Review Board: Biomedical Research and Innovation Center, Military Institute of Science and Technology, Mirpur -1216, Dhaka, Bangladesh
- Approval Status: Approved
- Approval Number: 2025H005
- Date: 20 Jun 2025
- Chairperson Signature: __________

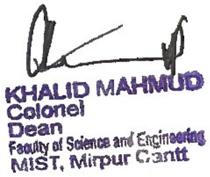
