## Supplementary material for "Comparative Evaluation of Mosquito Repellent Products in South Asia and North America: Efficacy, Safety, and Public Health Implications": Supplymentry materials: Ethical Approval Form Mosquito Repellent Study.pdf

### ETHICAL APPROVAL CERTIFICATE

#### Research Involving Human Volunteers

##### Title of the Study

---

#### Institutional Ethical Approval

This is to certify that the research proposal entitled **"Comparative Evaluation of Mosquito Repellent Products in South Asia and North America: Efficacy, Safety, and Public Health Implications"** submitted by the investigator(s) has been reviewed and approved by the Institutional Review Board: Biomedical Research and Innovation Center, Military Institute of Science and Technology, Mirpur -1216, Dhaka, Bangladesh in accordance with national and international ethical guidelines for research involving human participants.

The committee reviewed the objectives, methodology, volunteer recruitment procedures, informed consent process, safety precautions, risk minimization strategies, confidentiality measures, and overall ethical considerations associated with the study.

---

#### Study Information

**Research Title:**
Comparative Evaluation of Mosquito Repellent Products in South Asia and North America: Efficacy, Safety, and Public Health Implications

**Principal Investigator:**
________________________Prof Dr Md Ushama Shafoyat__________________

**Department/Institution:**
________________Military Institute of Science and Technology____________________

**Co-Investigator(s):**
__________________________________________

**Study Duration:**
_________________1 months_________________________

**Study Location:**
_______Dhaka North city corporation, Bangladesh___________________________

---

#### Human Volunteer Participation

A total of six (6) healthy adult volunteers participated in the study, including three males and three females aged between 18 and 22 years. The volunteers participated in:

- Arm-in-Cage Test Procedure
- Dermatological Safety Test

All participants were:

- Non-tobacco users
- Free from dermatosis or chronic skin disease
- Free from known allergic reactions to arthropod bites or mosquito repellents
- Informed regarding the objectives, procedures, risks, and benefits of the study

Written informed consent was obtained from all volunteers before participation.

Participants were instructed not to apply perfumes, aromatics, skincare products, or other mosquito repellents on the hands for at least 12 hours before and during the experiment to minimize experimental bias.

---

#### Safety and Ethical Considerations

1. All experiments were conducted under controlled laboratory conditions.
2. Appropriate safety precautions and monitoring procedures were maintained throughout the study.
3. Volunteers retained the right to withdraw from the study at any stage without penalty.
4. Confidentiality and anonymity of volunteer information were strictly maintained.
5. No severe physical or psychological risk was associated with participation.
6. Immediate medical support and first-aid facilities were available during experimentation.

---

#### Ethical Approval Statement

The Institutional Medical Ethics Committee has determined that the proposed research complies with ethical standards for studies involving human volunteers. The study protocol was approved after satisfactory review of the methodology, participant safety measures, and informed consent procedures.

##### Approval Information

**Approval Number/Code:** 2025H009

**Date of Approval:** ____12/ 09/2025____________________

**Validity of Approval:** __25/09/2025___________________

---

### Declaration

The investigators are responsible for conducting the study according to the approved protocol and ethical guidelines. Any modification to the study protocol must be reported to and approved by the Ethics Committee before implementation.

---

### Signatures

#### Principal Investigator

Name: _____Prof [illegible]yat_________

Signature: ______ ____________

Date: _____________25/09/2025_______________

---

#### Chairperson, Medical Ethics Committee

Name: Prof Khalid Mahmud, PhD

Designation: Dean, Faculty of Science and Technology

Institution: Military Institute of Science and Technology

Signature & Seal: ___________________________

KHALID MAHMUD
Colonel
Dean
Faculty of Science and Engineering
MIST, Mirpur Cantt

Date: ______25/09/2025__________________________
