## Supplementary material for "Comparative Evaluation of Mosquito Repellent Products in South Asia and North America: Efficacy, Safety, and Public Health Implications": Animal Ethical Approval

### ETHICAL APPROVAL APPLICATION FORMAT

**Animal Research (Rat Model)**

**1. Title of the Study**

**Safety and Toxicological Evaluation of Mosquito Repellent Products in a Rat Model**

**2. Principal Investigator (PI)**


- Name: Md Shafoyat
- Designation: Professor
- Department: Biomedical Enigneering
- Institution: Military Institute of Science and Technology
-


**4. Project Duration**

Start Date: 01 Jun 2025
End Date: 01 Jan 2026
Total Duration: 06 months

**5. Background and Rationale**

Mosquito repellent products are widely used for protection against vector-borne diseases such as dengue, malaria, and chikungunya. Despite their widespread use, limited studies have evaluated their systemic toxicity and safety profiles under controlled experimental conditions. This study aims to assess the potential toxicological effects of selected mosquito repellent formulations using a rat model to ensure safety for human exposure and guide public health recommendations.

**6. Objectives**

**General Objective:**

To evaluate the safety and toxicological effects of mosquito repellent products in a rat model.

**Specific Objectives:**

- To assess acute and sub-acute toxicity of selected mosquito repellents
- To evaluate hematological and biochemical changes in treated rats
- To observe histopathological changes in vital organs (liver, kidney, lung)
- To compare toxicity profiles between different repellent formulations

#### 7. Methodology

##### 7.1 Study Design

Experimental laboratory-based animal study.

##### 7.2 Animal Model

- Species: Albino Wistar rats (Rattus norvegicus)
- Sex: Male/Female (as applicable)
- Weight: 150–250 g
- Number of animals: 15

##### 7.3 Grouping of Animals

#### 10. Potential Risks

- Minimal risk to animals due to chemical exposure
- Proper monitoring and humane handling will reduce suffering

#### 11. Conflict of Interest

No conflict of interest

#### 12. Funding Source

- Self-funded

#### 13. Declaration by Principal Investigator

I hereby declare that the above information is correct and that the study will be conducted according to ethical guidelines for animal research. All necessary precautions will be taken to minimize [illegible]

Signature: _

Name: Md Shafoyat [illegible]

Date: 01 Jun 2025

#### 14. Ethical Review Committee Section

- Approval Status: Approved
- Approval Number: 2025H005
- Date: 20 Jun 2025

KHALID MAHMUD
Colonel
Dean
Faculty of Science and Engineering
MIST, Mirpur Cantt

- Chairperson Signature: ___________
